# Social isolation, loneliness, and blood-based AD/ADRD biomarkers in a nationally-representative study of middle-aged and older adults

**DOI:** 10.64898/2026.08.06.26359882

**Authors:** Karla Renata Flores Romero, Sirena Gutierrez, Scott C. Zimmerman, Anna M. Pederson, Mary Thoma, Ruijia Chen, Ashwin Kotwal, Maria Glymour, Kaitlin Casaletto, Jacqueline M. Torres

**Affiliations:** Department of Epidemiology and Biostatistics, University of California, San Francisco, San Francisco, CA; Department of Epidemiology, Boston University, Boston, MA; Department of Community Health Sciences, UCLA Fielding School of Public Health, Los Angeles, CA; Division of Geriatrics, School of Medicine, University of California, San Francisco, CA; Department of Neurology, University of California, San Francisco, CA; Memory and Aging Center, University of California, San Francisco, CA

## Abstract

**Importance:** The biological mechanisms underlying the associations of social isolation and loneliness with dementia risk are not well understood.

**Objective:** To evaluate the relationship of prospectively measured social isolation and loneliness with AD/ADRD blood-based biomarkers.

**Design:** Observational study using the U.S. Health and Retirement Study (2010-2016). Venous blood draws were conducted in 2016 and AD/ADRD biomarkers were released in 2025. We estimated associations of social isolation and loneliness patterns between 2012 and 2014 with continuous biomarkers using linear regressions, accounting for socio-demographic and health covariates. We evaluated effect modification by sex and APOE ɛ4 carrier status.

**Setting:** Population-based

**Participants:** Community-dwelling HRS participants aged 50 years or older (n = 3862).

**Exposures:** Primary exposures were four-category multi-wave variables of persistent, resolving, new-onset, or no social isolation/loneliness across the two exposure waves. Social isolation was classified as “severe” and “moderate-to-severe” based on a 5-item scale including marital status, household size, proximity to children, religious service attendance, and volunteering. Past-week loneliness was measured with a single-item question (yes/no).

**Main Outcomes and Measures:** Neurofilament light chain (NfL), glial fibrillary acidic protein (GFAP), and the ratio of amyloid beta 42 to amyloid beta 40 (Aβ42/40), measured in plasma via a Multiplex Simoa Assay and phosphorylated tau (p-tau181), measured in serum via a Simoa Assay.

**Results:** At the analytic baseline, respondents were a mean age of 64 (9.5) years, 59% female, and 25% APOE ɛ4 carriers. Across the two exposure waves, 4% experienced persistent severe social isolation, 16% experienced persistent moderate-to-severe social isolation, and 8% reported persistent loneliness. Multiple patterns of social isolation (vs. no social isolation) were associated with higher NfL, including persistent severe social isolation (*β*: 0.31), new-onset moderate-to-severe social isolation (*β*: 0.14), and resolving moderate-to-severe social isolation (*β*: 0.20). Persistent severe social isolation was associated with lower GFAP (*β*: − 0.31) while persistent loneliness and, for men, new-onset loneliness were associated with higher GFAP (*β_persistent_*: 0.16; *β_new_*_*onset*_*men*_: 0.25). New-onset severe social isolation was associated with a lower Aβ42/40 ratio (*β*: − 0.25) while resolving moderate-to-severe social isolation and, for men, persistent severe social isolation were each associated with higher p-tau181 (*β_resolving_*: 0.11; *β_persistent_*__*men*_: 0.41). There was some additional variation by APOE ɛ4 carriership, although selective survival is a concern.

**Conclusions:** Social isolation was associated with elevated blood-based biomarkers of neuronal injury, with variation by patterns of exposure over time. Associations between social isolation and loneliness with biomarkers related to astrocyte damage and Alzheimer’s disease were less consistent, and varied in sign and magnitude by exposure and sex.

## Introduction

Social isolation and loneliness are modifiable risk factors associated with cognitive decline and risk of Alzheimer’s disease and related dementias (AD/ADRD).^1–5^ However, evidence linking these social exposures to the biological mechanisms underlying dementia risk (i.e. how social exposures get “under the skin”) is limited.^2–6^ Evaluating associations of social isolation and loneliness with biomarkers of dementia-related biological pathways may improve understanding of the mechanisms linking these social exposures to dementia, provide insights into disease etiology, and inform the development of biomarkers for risk and/or monitoring the efficacy of social isolation and loneliness interventions.

Prior studies evaluating the associations of social isolation and loneliness with structural^7–10^ and functional^11–15^ neuroimaging or fluid biomarker-based outcomes relevant to AD/ADRD have yielded mixed evidence.^16–18^ These findings have been drawn on heterogenous data sources, including the UK biobank,^19^ the Baltimore Longitudinal Study on Aging, the Framingham Heart Study,^20^ the Harvard Aging Brain Study,^16^ and others.^21^ However, these studies were based on clinical or convenience samples that limited generalizability, cross-sectional designs that exacerbated concerns about reverse causation,^22^ and, in many cases, fairly small samples resulting in imprecise estimates.

Blood-based AD/ADRD biomarkers provide a scalable and minimally invasive means of capturing underlying biological processes in large cohorts. These include biomarkers that reflect Alzheimer’s disease-specific pathology, such as markers of amyloid or tau pathology, as well as more general markers of neuronal injury and astrocytic damage that are elevated in the context of multiple neurodegenerative diseases. The feasibility of these biomarkers enables evaluation of associations in large, geographically dispersed, population-based samples. We examined the associations of prospectively measured social isolation and loneliness with AD/ADRD blood-based biomarkers in a large, population-based study of older adults in the U.S. We further evaluated effect modification by sex and apolipoprotein E ε4 (APOE ε4) carrier status given prior evidence of heterogeneous associations between social isolation, loneliness, and dementia risk by these factors.^23–25^

## Methods

### Data

We used data from the U.S. Health and Retirement Study (HRS), a nationally representative longitudinal study of community-dwelling adults aged 50 years and older and their spouses or partners of any age with biennial follow-up. In 2016, the HRS fielded a Venous Blood Study (VBS), in which eligible respondents who completed the 2016 HRS interview were invited to complete an in-home venous blood draw.^26^ A total of 9,932 participants completed the VBS. From these participants, a probability sample of stored biospecimens (n = 4539) was selected for blood-based AD/ADRD biomarker analyses, with biomarker data publicly released in December 2025. Characteristics of VBS participants and nonparticipants are presented in Appendix Table 1.

**Table 1.** Baseline (2010) descriptive characteristics of HRS respondents 50+ participating in biennial waves from 2010 to 2016 and with blood-based AD/ADRD biomarker data.

|  | **Unweighted**<br>**(n=3862)** |
| --- | --- |
| **Baseline covariates** |  |
| Age, mean (SD) | 62.7 (9.2) |
| Female, No. (%) | 2,274 (54.6%) |
| **Race/Ethnicity, No. (%)** |  |
| Non-Hispanic White | 2,581 (78.8%) |
| Non-Hispanic Black | 638 (9.4%) |
| Hispanic | 537 (8.9%) |
| Other | 106 (2.9%) |
| Educational attainment (yrs.) | 13.3 (3.0) |
| **Father's educational attainment, No. (%)** |  |
| ≤ mean | 1,464 (34.2%) |
| > mean | 1,854 (54.0%) |
| Missing | 544 (11.9%) |
| **Mother's educational attainment, No. (%)** |  |
| ≤ mean | 1,372 (30.4%) |
| > mean | 2,197 (63.1%) |
| Missing | 293 (6.5%) |
| **Marital status, No. (%)** |  |
| Married/Partnered | 2,662 (69.4%) |
| Divorced/Separated | 530 (13.7%) |
| Widowed | 468 (10.0%) |
| Never married | 202 (6.8%) |
| **Spouse's educational attainment, No. (%)** |  |
| ≤ mean | 1,281 (29.6%) |
| > mean | 1,335 (38.7%) |
| Missing | 1,246 (31.7%) |
| **Time-varying covariates** |  |
| **Chronic health conditions, No. (%)** |  |
| 0 conditions | 670 (19.2%) |
| 1-2 conditions | 2,065 (53.4%) |
| ≥3 conditions | 1,127 (27.4%) |
| **Self-rated hearing** |  |
| Excellent or very good | 1,851 (50.2%) |
| Good | 1,324 (32.6%) |
| Fair or Poor | 686 (17.2%) |
| Missing | 1 (0.0%) |
| **Self-rated vision** |  |
| Excellent or very good | 1,328 (37.7%) |
| Good | 1,699 (42.1%) |
| Fair or Poor | 829 (20.1%) |
| Missing | 6 (0.2%) |
| **Cognitive status, No. (%)** |  |
| Cognitive Impairment, Not Dementia | 477 (9.9%) |
| Dementia | 66 (1.5%) |
| Current smoking, No. (%) | 557 (13.2%) |
| Missing | 17 (0.3%) |
| Current Drinking, , No. (%) | 2,379 (64.7%) |
| Any weekly physical activity, No. (%) | 3,560 (92.6%) |
| Missing | 190 (4.8%) |
| Elevated depression, No. (%) | 292 (7.0%) |
| Source: Health and Retirement Study (HRS), 2010-2016. Values are presented as unweighted counts and weighted percentages. Means and percentages were estimated using the 2016 VBS innovative sample weights. A complete version of this table, including unweighted estimates and estimates weighted using both the 2016 VBS full sample weights and innovative sample weights, is provided in Appendix Table 5. |  |

Our analytic sample included community-dwelling HRS respondents aged 50 years or older at the 2010 analytic baseline who had complete exposure, covariate, and 2016 AD/ADRD biomarker data (n = 3862, Appendix Figure 1). To align with our hypothesized temporal ordering (see Directed Acyclic Graph, Appendix Figure 2), we drew exposure data from the 2012 and 2014 HRS waves (i.e., two and four years prior to outcome assessment). Covariates were measured two years before each exposure assessment (i.e. 2010 and 2012, respectively). This study followed STROBE reporting guidelines for observational research.^27^

**Figure 1.**
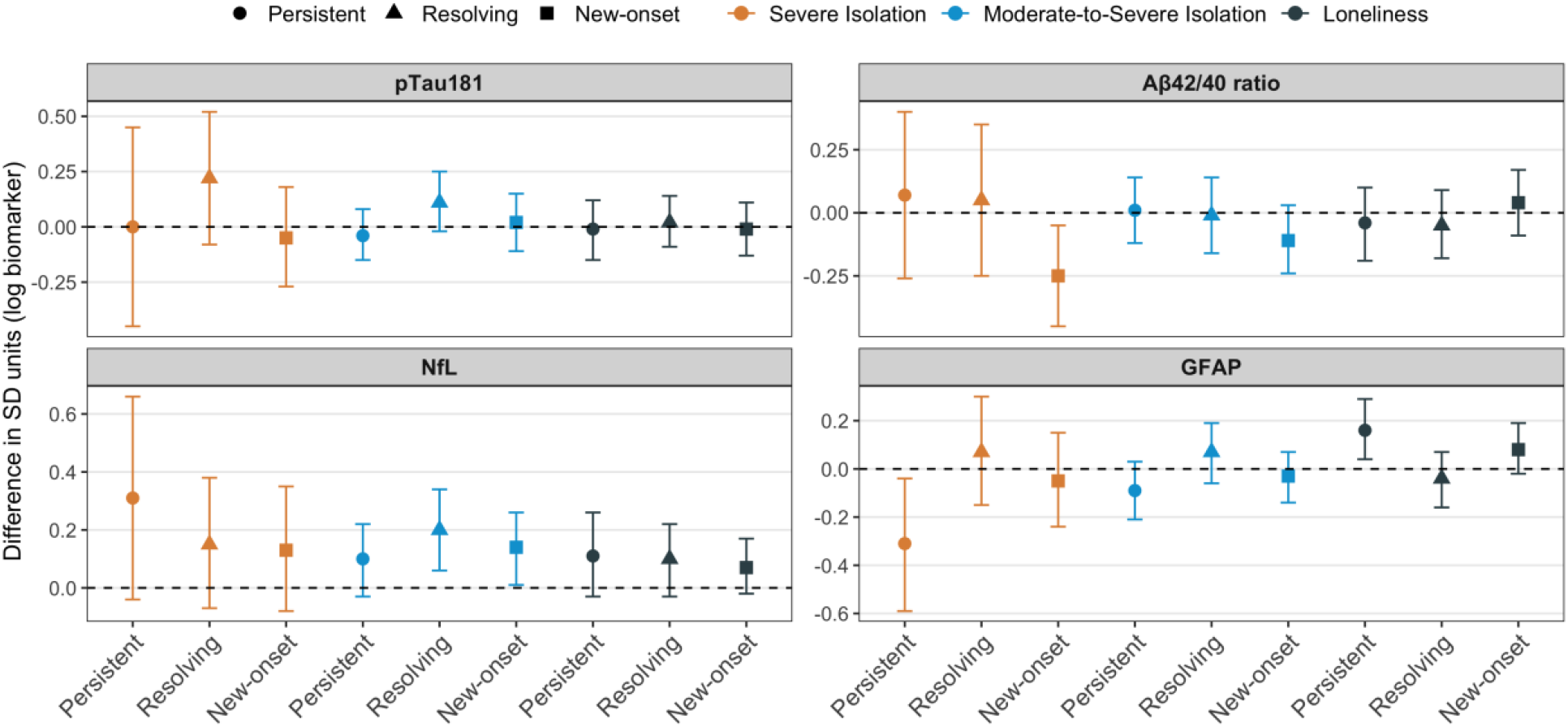
Associations between four-category multi-wave isolation and loneliness exposure variables and AD/ADRD biomarkers in the U.S. Health and Retirement Study. Data are from the U.S. Health and Retirement Study (HRS), 2010–2016 (n=3862). Points represent estimated differences in standard deviation (SD) units of log-transformed biomarker levels, and error bars indicate 95% confidence intervals. Four-category multi-wave variables for severe isolation (<1 point), moderate-to-severe isolation (<2 points), and loneliness were constructed using assessments from 2012 and 2014 to classify participants as having no exposure at either wave (reference), exposure at both waves (persistent), exposure in 2012 but not 2014 (resolving), or exposure in 2014 both not 2012 (new onset). Estimates are based on linear regression models with stabilized inverse probability of treatment and retention weights and robust standard errors. Models adjust for baseline covariates (age, sex, race/ethnicity, and educational attainment).

**Figure 2.**
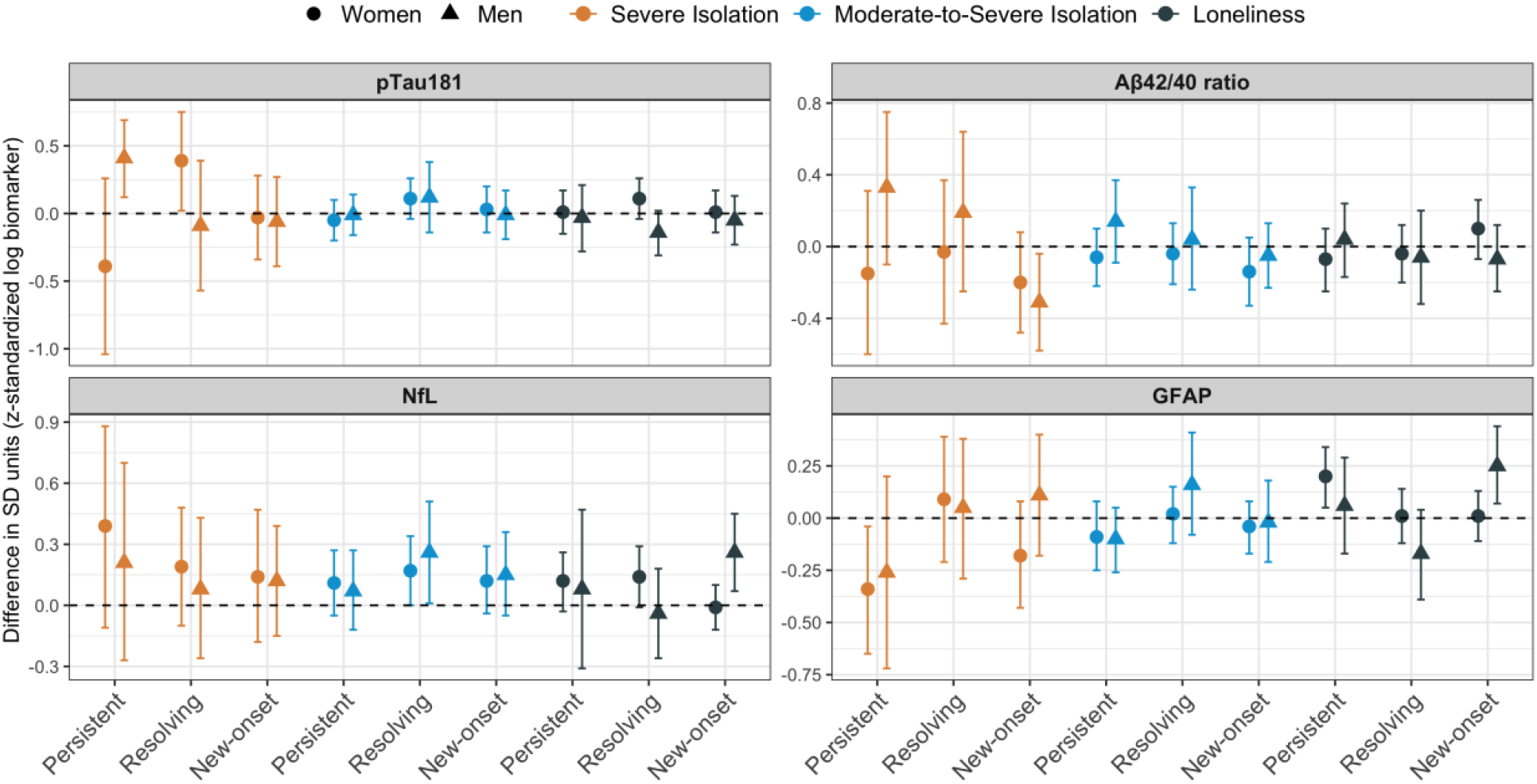
Associations between four-category multi-wave isolation and loneliness exposure variables and AD/ADRD biomarkers, Stratified by Sex, in the U.S. Health and Retirement Study. Data are from the U.S. Health and Retirement Study (HRS), 2010–2016; (Women: n = 2274; Men: n = 1588). Points represent estimated differences in standard deviation (SD) units of log-transformed biomarker levels, and error bars indicate 95% confidence intervals. Four-category multi-wave variables for severe isolation (<1 point), moderate-to-severe isolation (<2 points), and loneliness were constructed using assessments from 2012 and 2014 to classify participants as having no exposure at either wave (reference), exposure at both waves (persistent), exposure in 2012 but not 2014 (resolving), or exposure in 2014 both not 2012 (new onset). Estimates are based on linear regression models with stabilized inverse probability of treatment and retention weights and robust standard errors. Models adjust for baseline covariates (age, sex, race/ethnicity, and educational attainment). P values for interactions: severe isolation: persistent (p-tau181 = .02, Aβ42/40 = .16, NfL = .60, GFAP = .79), resolving (p-tau181 = .13, Aβ42/40 = .48, NfL = .62, GFAP = .84), and new-onset (p-tau181 = .89, Aβ42/40 = .57, NfL = .91, GFAP = .14); moderate-to-severe isolation: persistent (p-tau181 = .55, Aβ42/40 = .23, NfL = .65, GFAP = .76), resolving (p-tau181 = .89, Aβ42/40 = .69, NfL = .63, GFAP = .39), and new-onset (p-tau181 = .82, Aβ42/40 = .61, NfL = .85, GFAP = .98); loneliness: persistent (p-tau181 = .99, Aβ42/40 = .54, NfL = .85, GFAP = .25), resolving (p-tau181 = .05, Aβ42/40 = .80, NfL = .19, GFAP = .10), and new-onset (p-tau181 = .73, Aβ42/40 = .17, NfL = .02, GFAP = .03).

## Measures

### Exposure

Social isolation was measured in 2012 and 2014, two and four years prior to outcome assessment, using a modified version of a scale developed by Cenzer et al^28^ based on items from the HRS Core interview. The measure includes five items: marital status, household size, proximity to children, frequency of religious service attendance, and participation in volunteer activities (Appendix Table 2). Total scores range from 0 to 5, with lower scores indicating greater isolation. Social isolation was defined using two binary thresholds: severe isolation (< 1 point) and moderate-to-severe isolation (< 2 points). For each threshold, we created a four-category multi-wave exposure variable representing persistent (present at both waves), new-onset (present only in 2014), resolving (present only in 2012), or no social isolation across the two exposure waves. These multi-wave exposure variables were the primary exposures in the main analyses.

Loneliness was measured in 2012 and 2014 using a single-item asking whether respondents felt lonely “much of the time” during the past week (yes/no). This item, which is included within a modified version of the 8-item Centers for Epidemiologic Studies – Depression (CES-D) scale,^29,30^ corresponds closely to cut-offs used in longer and well-validated loneliness scales.^31^ We similarly created a four-category multi-wave exposure variable representing persistent, new-onset, resolving or no loneliness across the two exposure waves. This multi-wave exposure variable was the primary loneliness exposure in the main analyses.

### Outcomes

Blood samples were collected from consenting respondents following the 2016 HRS interview and stored samples were processed using standardized protocols at the University of Minnesota Advanced Research and Diagnostic Laboratory.^26^ Phosphorylated tau at threonine 181 (p-tau181), a marker of tau pathology associated with Alzheimer’s disease, was measured in serum using a Simoa Assay (Quanterix, MA). The ratio of amyloid beta 42 to amyloid beta 40 (Aβ42/40), a marker of cerebral amyloid pathology associated with Alzheimer’s disease, and neurofilament light chain (NfL) and glial fibrillary acidic protein (GFAP), markers of neuroaxonal injury and astrocytic activation, respectively, were measured in plasma using a Multiplex Simoa Neurology 4-Plex assay (Quanterix, MA).

### Covariates

Baseline covariates included sociodemographic characteristics (age, sex, race and ethnicity), marital status (included in models of loneliness only), and lifecourse socioeconomic status, including respondent educational attainment, parental educational attainment, and spouse/partners educational attainment, with an indicator for respondents who were not married or partnered at analytic baseline.

Time-varying covariates were measured two years before exposure assessment and included a count of self-reported physician-diagnosed chronic conditions (hypertension, diabetes, cancer, lung disease, heart disease, stroke, psychiatric condition, and arthritis) as a measure of overall chronic disease burden, an indicator of probable cognitive impairment or dementia status based on the Langa-Weir algorithm to minimize reverse confounding,^32^ and separate indicators of self-rated hearing and vision. Additional time-varying covariates included current smoking, current alcohol use, physical activity, and past-week depressive symptoms; these variables were included in sensitivity analyses only. Depressive symptoms were measured using the 8-item Centers for Epidemiologic Studies – Depression scale,^29,30^ excluding the loneliness item.

### Effect Modifiers

We evaluated effect modification by binary indicators of respondent sex in our main analytic sample and by *APOE* ɛ4 carrier status in a subset of participants with available genotype data (n = 3546).

## Statistical Analysis

We first estimated unweighted and weighted descriptive statistics, focusing on covariates measured at analytic baseline, the primary four-category social isolation and loneliness exposure measures, and AD/ADRD biomarker values measured at six-year follow-up. See Appendix Text 1 for further details on the HRS sample weights used.

We then estimated the association between the multi-wave social isolation and loneliness exposure variables (with no exposure at either wave as the reference group) and AD/ADRD biomarkers using marginal structural models with inverse probability of treatment weights (IPTWs) to account for time-varying covariates.^33,34^ This approach appropriately accounts for time-varying confounders that may be influenced by prior exposure by creating a weighted pseudo-population in which measured confounders are balanced across exposure groups. To account for attrition between 2010 analytic baseline and 2016 outcome assessment, we additionally constructed inverse probability of retention weights (IPRWs). The product of the IPTWs and IPRWs was applied to linear regression models with robust standard errors to estimate the association between each exposure and biomarker outcome. Further details on the construction of the IPTWs and IPRWs are provided in Appendix Text 2 and Appendix Table 3.

We used a complete-case approach for age, sex, race and ethnicity, respondent’s own educational attainment, marital status, spouse/partner educational attainment, chronic health conditions, and probable cognitive impairment or dementia status. Missing values for all remaining covariates were modeled using a separate indicator category (Appendix Table 4).

Because biomarker distributions were right-skewed, our main analyses used natural log-transformed biomarker values, which were subsequently standardized to facilitate comparison of effect estimates across biomarkers. We also conducted supplementary analyses using (1) log-transformed biomarkers without standardization, and (2) standardized biomarkers without log transformation. For the supplementary analyses using log-transformed outcomes, coefficients and 95% confidence intervals were exponentiated and expressed as percent differences to facilitate interpretation.

We evaluated effect modification separately by respondent sex and by APOE ε4 carrier status via multiplicative interaction terms and stratified analyses.

Sensitivity analyses evaluated the robustness of our findings to: 1) incorporation of HRS weights (Appendix Text 1), 2) additional adjustment for lagged time-varying health behaviors and depressive symptoms (Appendix Text 3); and 3) additional adjustment for APOE ε4 carrier status among participants with available genotype data (Appendix Text 4).

As secondary analyses, we alternatively estimated associations using single-time-point measures of social isolation and loneliness assessed in 2012 and 2014 (at two and four years before outcome assessment) rather than the primary multi-wave exposure variables.

All analyses were conducted in STATA 18.5.

## Results

At the analytic baseline (2010), mean respondent age was 64 years (± 9.5 SD), 59% of participants were women, 79% were non-Hispanic White, 9% were non-Hispanic Black, 9% were Hispanic, and 25% of participants were *APOE* ε4 carriers (Table 1; full descriptive statistics are in Appendix Table 5).

Exposure status was relatively stable across the two exposure waves (Appendix Figure 3). Four percent of participants were classified as experiencing persistent severe isolation, 2% as experiencing resolving severe isolation, and 2% as experiencing new-onset severe isolation. Corresponding prevalences for moderate-to-severe isolation were 16% (persistent), 6% (resolving), and 7% (new-onset). Eight percent of participants reported persistent loneliness, 7% reported resolving loneliness, and 8% reported new-onset loneliness (Appendix Table 6). Median (IQR) biomarker concentrations measured in 2016 were 1.6 pg/mL for p-tau181 (1.1– 2.5), 0.1 for the Aβ42/40 ratio (0.1-0.1), 18 pg/mL for NfL (11.9–29.2), and 87.7 pg/mL for GFAP (56.7–134.5) (Appendix Table 7).

**Figure 3.**
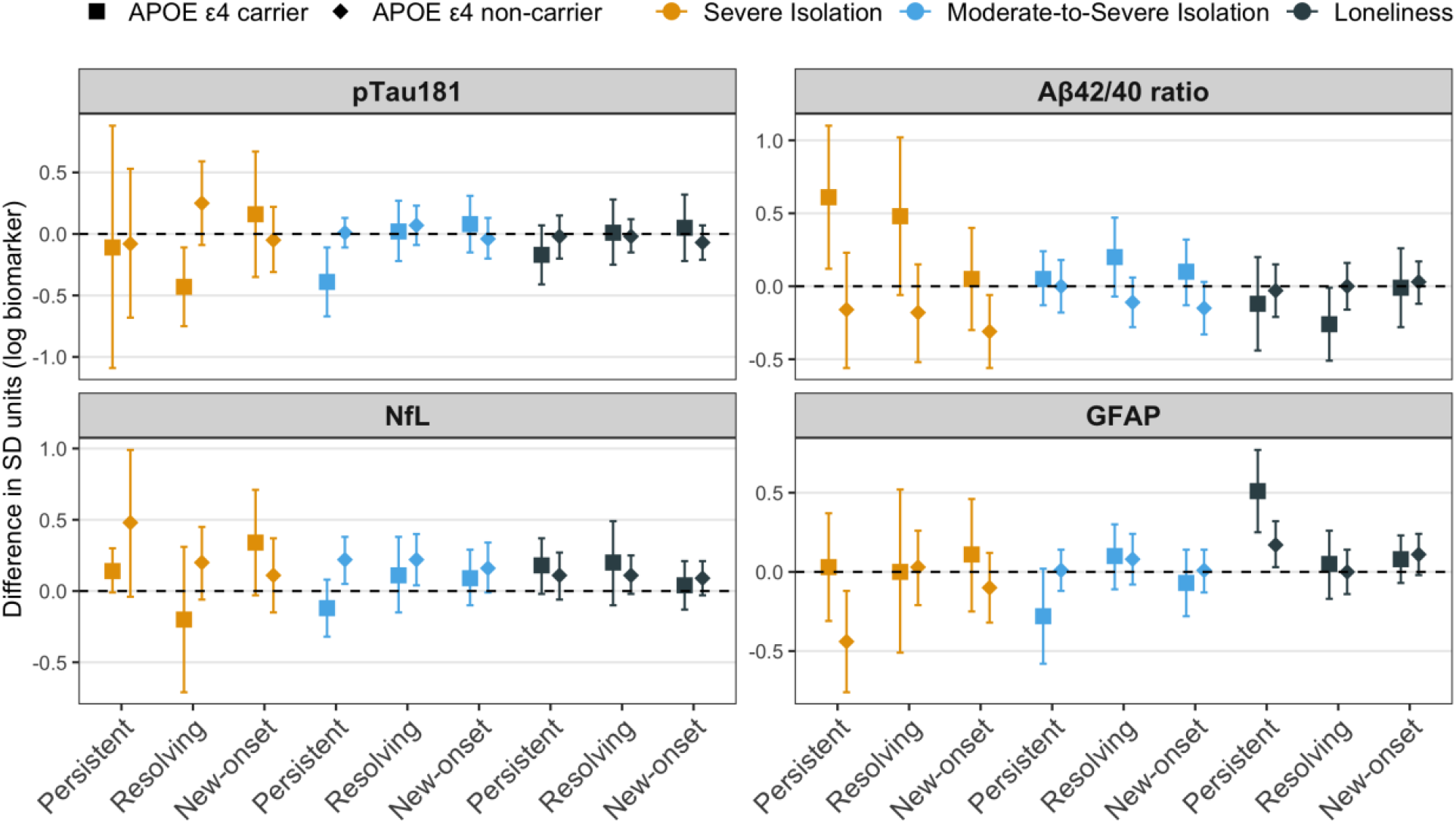
Associations between four-category multi-wave isolation and loneliness exposure variables and AD/ADRD biomarkers, Stratified by APOE ε4 carrier status, in the U.S. Health and Retirement Study. Data are from the U.S. Health and Retirement Study (HRS), 2010–2016; (APOE ε4 carrier: n = 903; Non-APOE ε4 carrier: n = 2,643). Points represent estimated differences in standard deviation (SD) units of log-transformed biomarker levels, and error bars indicate 95% confidence intervals. Four-category multi-wave variables for severe isolation (<1 point), moderate-to-severe isolation (<2 points), and loneliness were constructed using assessments from 2012 and 2014 to classify participants as having no exposure at either wave (reference), exposure at both waves (persistent), exposure in 2012 but not 2014 (resolving), or exposure in 2014 both not 2012 (new-onset). Estimates are based on linear regression models with stabilized inverse probability of treatment and retention weights and robust standard errors. Models adjust for baseline covariates (age, sex, race/ethnicity, and educational attainment. P values for interactions: severe isolation: persistent (p-tau181 = .97, Aβ42/40 = .01, NfL = .25, GFAP = .03), resolving (p-tau181 = .009, Aβ42/40 = .05, NfL = .18, GFAP = .96), and new-onset (p-tau181 = .44, Aβ42/40 = .06, NfL = .31, GFAP = .28); moderate-to-severe isolation: persistent (p-tau181 = .03, Aβ42/40 = .46, NfL = .03, GFAP = .27), resolving (p-tau181 = .84, Aβ42/40 = .05, NfL = .62, GFAP = .65), and new-onset (p-tau181 = .41, Aβ42/40 = .10, NfL = .62, GFAP = .58); loneliness: persistent (p-tau181 = .21, Aβ42/40 = .82, NfL = .82, GFAP = .04), resolving (p-tau181 = .96, Aβ42/40 = .09, NfL = .82, GFAP = .99), and new-onset (p-tau181 = .50, Aβ42/40 = .97, NfL = .49, GFAP = .84).

### Social isolation, loneliness, and blood-based AD/ADRD biomarkers

Persistent (vs. no exposure) severe isolation was associated with higher NfL (*β*: 0.31, 95% Confidence Interval [CI]: −0.04 to 0.66) and lower GFAP (*β*: − 0.31, 95% CI: −0.59 to −0.04) (Figure 1; Appendix Table 8). New-onset severe isolation was associated with a lower Aβ42/40 ratio (*β*: − 0.25, 95% CI: −0.45 to −0.05). Resolving moderate-to-severe isolation was associated with higher p-tau181 (*β*: 0 .11, 95% CI: −0.02 to 0.25) and higher NfL (*β*: 0.20, 95% CI: 0.06 to 0.34). New-onset moderate-to-severe isolation was associated with higher NfL (*β*: 0.14, 95% CI: 0.01 to 0.26). Persistent loneliness was associated with higher GFAP (*β*: 0.16, 95% CI: 0.04 to 0.29).

Conclusions using alternative outcome specifications (i.e., log-transformed biomarkers expressed as percent differences and standardized biomarkers) were generally similar to those based on standardized log-transformed biomarkers, although some associations differed in magnitude and statistical significance in the standardized models (Appendix Tables 9 and 10).

### Effect modification by sex

Persistent severe isolation was associated with higher p-tau181 among men (β: 0.41, 95% CI: 0.12 to 0.69) but not among women (P for interaction = .02; Figure 2; Appendix Table 11). Resolving loneliness was associated with lower p-tau181 among men (β: −0.14, 95% CI: −0.31 to 0.02) but not among women (P for interaction = .05). New-onset loneliness was associated with higher NfL among men (β: 0.26, 95% CI: 0.07 to 0.45) and higher GFAP among men (β: 0.25, 95% CI: 0.07 to 0.44) but not among women (P for interaction = .02 and .03, respectively).

### Effect modification by APOE ε4 carrier status

Among APOE ε4 carriers, we found some counterintuitive associations between social isolation measures and more favorable AD/ADRD biomarker values. For example, among APOE ε4 carriers, persistent moderate-to-severe isolation was associated with lower p-tau181 (β: −0.39, 95% CI: −0.67 to −0.11), whereas no association was observed among non-carriers (P for interaction = .03) (Figure 3; Appendix Table 12). Conversely, among non-carriers, persistent moderate-to-severe isolation was associated with higher NfL (β: 0.22, 95% CI: 0.05 to 0.38), whereas no association was observed among APOE ε4 carriers (P for interaction = .03).

In another example, the association between persistent loneliness and GFAP was stronger among APOE ε4 carriers (β: 0.51, 95% CI: 0.25 to 0.77) than among non-carriers (β: 0.17, 95% CI: 0.03 to 0.32) (P for interaction = .04).

### Sensitivity analyses

Results were broadly consistent with the main unweighted analyses after applying HRS weights, although several associations were attenuated, including those of new-onset severe isolation with Aβ42/40 ratio and of persistent loneliness with GFAP, which were no longer statistically significant (Appendix Figure 4 and Appendix Table 13). Similarly, results were generally consistent with the main analyses after additional adjustment for lagged time-varying health behaviors and depressive symptoms (Appendix Table 14). Finally, models that additionally adjusted for APOE ε4 carrier status among a subset with APOE genotype were generally similar to the main analyses, although the magnitude of several estimates was larger and in some cases became statistically significant (Appendix Table 15).

We alternatively estimated associations using single-time-point measures of social isolation and loneliness at two and four years before outcome assessment (see Appendix Tables 16 for descriptive statistics and Appendix Figure 5 and Appendix Table 17 for estimates). Severe isolation measured at two years before outcome assessment was associated with a lower Aβ42/40 ratio (*β*: −0.22: 95% CI: −0.40 to −0.03). Moderate-to-severe isolation measured four years before outcome assessment was associated with higher NfL (*β*: 0.15, 95% CI: 0.03 to 0.27), whereas loneliness measured four years before outcome assessment was associated with higher GFAP values (*β*: 0.11, 95% CI: 0.02 to 0.19).

## Discussion

In this cohort study, we found evidence that social isolation and loneliness were associated with blood-based AD/ADRD biomarkers, although associations varied by exposure pattern and type, biomarker outcome, and participant sex, and some associations were attenuated under alternative model specifications.The most consistent evidence pointed to associations between social isolation and blood-based measures of neurofilament light chain (NfL), a protein that is elevated in response to neuronal injury. There was some evidence of association between persistent loneliness and glial fibrillary acidic protein (GFAP), which is elevated in the presence of astrocyte damage. However, estimates in the overall sample were not robust to alternative modeling specifications, and the magnitude varied by APOE ε4 carrier status. We also note that we observed associations between persistent severe isolation and *lower* GFAP, complicating clear conclusions. Associations with biomarkers specific to Alzheimer’s disease processes, p-tau181 and Aβ42/40 ratio, were less common. To our knowledge, this is the first population-based study to evaluate prospective associations of social isolation and loneliness with blood-based AD/ADRD biomarkers.

Our results generally suggest an association between social isolation and a blood-based marker linked to neuroaxonal injury. While these associations varied in magnitude and precision according to exposure pattern (persistent, resolving, and new-onset) and social isolation specification (severe or moderate-to-severe), they remained generally consistent across alternative model specifications. These associations could be driven by upstream influences of social isolation on adverse health behaviors (e.g., smoking and physical inactivity^35^), poorer psychological wellbeing (e.g., elevated depressive symptoms^36^), and adverse physiological outcomes^37^ (e.g., hypothalamic-pituitary-adrenal axis dysregulation,^38^ inflammation,^39^ disrupted sleep^40^). These behavioral, psychological, and physiological factors are well-established risk factors for a range of clinical neurodegenerative outcomes, although evidence linking these factors to blood-based AD/ADRD biomarkers remains limited.^41,42^

Associations between loneliness and blood-based AD/ADRD biomarkers were less consistent, although we found important variation by sex. Among men, but not women, new-onset loneliness was associated with higher NfL and GFAP. This is in contrast to analyses of UK Biobank and Baltimore Longitudinal Study of Aging data, which found no association between loneliness, including changes in loneliness, and blood-based AD/ADRD biomarkers, nor evidence of effect modification by sex.^19^ However, prior studies have reported sex differences in the relationship between loneliness and dementia risk, including evidence that new-onset loneliness is associated with increased dementia risk among men.^43,44^ These differences may reflect variation in the specific dimensions of loneliness experienced (i.e., intimate, relational, and collective).^45,46^ Further research is needed to clarify the mechanisms underlying these sex differences.

We also observed evidence that the magnitude and direction of some associations, particularly those involving social isolation, differed by APOE ε4 carrier status, although confidence intervals overlapped across stratified analyses. The number of APOE ε4 carriers who were also socially isolated and/or lonely was relatively small, resulting in imprecise estimates. Selection processes may have influenced these findings. APOE ε4 carriers are at increased risk of mortality,^47^ cognitive decline,^48^ and study attrition,^49^ and those who survived and participated in biomarker collection may represent a particularly healthy subset of carriers. Consequently, survival and selection bias may operate at the intersection of APOE ε4 carrier status and social isolation, complicating interpretation of apparent heterogeneity.^50^ Moreover, because early neurodegenerative processes may contribute to changes in social relationships,^10^ reverse causation may be particularly difficult to disentangle among APOE ε4 carriers, who are at elevated risk of cognitive decline. Together, selective survival, differential participation, reverse causation, and residual confounding may help explain why seemingly protective associations between social isolation and AD/ADRD biomarkers were observed among ε4 carriers.

Several additional limitations should be noted. First, although we adjusted for a range of sociodemographic and health-related covariates, residual confounding remains possible. Second, there are limitations related to the AD/ADRD biomarkers available in the HRS.^51^ Biomarkers were assessed at a single time-point, limiting our ability to examine longitudinal changes or account for baseline biomarker levels to better address potential reverse causation. In addition, research suggests that p-tau217, which was not available, is superior to p-tau181 at predicting amyloid positivity as measured by positron emission tomography.^52^ There are also known drawbacks to using the Simoa platform to measure Aβ42/40.^53^

Fourth, although findings were generally robust across alternative model specifications and additional covariate adjustment, several associations were attenuated after applying HRS weights, suggesting that some estimates may be sensitive to sample selection and weighting assumptions. Finally, we were unable to use the more comprehensive measures of social isolation and loneliness collected as part of the HRS leave-behind questionnaire, which captures multiple dimensions of both constructs.^46^ The subset of participants with both leave-behind questionnaire data and blood-based AD/ADRD biomarker measurements was too small to support adequately powered subgroup analyses.

## Conclusion

In this cohort study of middle-aged and older adults in the US, social isolation and loneliness were prospectively associated with blood-based biomarkers of neuronal injury and astrocyte damage, albeit with complex heterogeneity by exposure pattern, sex, and model specification. Social isolation was most consistently associated with higher NfL levels, with some variation across exposure specifications (i.e., severe or moderate-to-severe) and longitudinal exposure patterns. We observed some evidence of association between some specifications of loneliness and higher levels of NFL and GFAP for men, in particular, although we also observed counterintuitive associations between persistent social isolation and higher levels of GFAP, complicating clear conclusions. These findings suggest that social isolation and loneliness may contribute to neurodegenerative processes through distinct biological pathways, underscoring the potential importance of social relationships for brain health in later life. These are, to our knowledge, the first national-level, population-based estimates of prospective associations between social isolation, loneliness, and blood-based AD/ADRD biomarkers. However, these warrant replication in other populations and extension to a broader set of biomarkers as well as longitudinal biomarker measures.

## Question

What are the relationships between social isolation and loneliness with blood-based biomarkers relevant to Alzheimer’s disease and related dementias in a national, population-based study of adults in the United States?

## Findings

Social isolation and loneliness were associated with blood-based biomarkers of neuronal injury and astrocyte damage, with variation by how isolation/loneliness changed over time and by sex. Associations with Alzheimer’s-specific biomarkers were limited, and apparent differences by APOE ε4 status were inconsistent and may reflect selective survival rather than true biological effect modification.

## Meaning

Our findings suggest potential biological mechanisms, primarily related to general neural and glial degeneration, that may explain prior associations of social isolation and loneliness with dementia risk.

## Data Availability

All data produced in the present study are available upon reasonable request to the authors.

https://hrs.isr.umich.edu/data-products

