## Supplementary Appendix for "Social isolation, loneliness, and blood-based AD/ADRD biomarkers in a nationally-representative study of middle-aged and older adults"

#### Appendix Tables and Figures

##### eTable 1. Comparison of Characteristics of Eligible Participants Who Did and Did Not Participate in the 2016 Venous Blood Study

| | Eligible Nonparticipants (n = 5577) | Participants (n = 9932) | P value[1] |
|---|---|---|---|
| **Covariates** | | | |
| Age, mean (SD) | 68.6 (10.7) | 68.6 (10.2) | 0.741 |
| Female, n (%) | 3412 (61.2%) | 5866 (59.1%) | 0.010 |
| Race/Ethnicity, n (%) | | | |
| Non-Hispanic White | 3144 (56.5%) | 6379 (64.3%) | <0.001 |
| Non-Hispanic Black | 1336 (24.0%) | 1747 (17.6%) | |
| Hispanic | 864 (15.5%) | 1476 (14.9%) | |
| Other | 221 (4.0%) | 313 (3.2%) | |
| Educational attainment (yrs) | 12.8 (3.2) | 12.8 (3.2) | 0.285 |
| Marital status, n (%) | | | |
| Married/Partnered | 3180 (57.2%) | 6380 (64.3%) | <0.001 |
| Divorced/Separated | 893 (16.0%) | 1350 (13.6%) | |
| Widowed | 1155 (20.8%) | 1750 (17.6%) | |
| Never married | 336 (6.0%) | 440 (4.4%) | |
| Chronic health conditions , n (%) | | | |
| 0 conditions | 718 (12.9%) | 1046 (10.5%) | <0.001 |
| 1-2 conditions | 2584 (46.3%) | 4434 (44.6%) | |
| ≥3 conditions | 2275 (40.8%) | 4452 (44.8%) | |
| Cognitive status, n (%) | | | |
| CIND | 1094 (19.6%) | 1718 (17.3%) | <0.001 |
| Dementia | 322 (5.8%) | 393 (4.0%) | |
| Current smoking, n (%) | 757 (13.6%) | 1152 (11.6%) | 0.001 |
| Missing | 26 (0.5%) | 52 (0.5%) | |
| Current Drinking, , n (%) | 3018 (54.1%) | 5574 (56.1%) | 0.017 |
| Any weekly physical activity, n (%) | 4821 (86.4%) | 8814 (88.7%) | <0.001 |
| Missing | 351 (6.3%) | 560 (5.6%) | |
| Elevated depression, n (%) | 593 (10.6%) | 1020 (10.3%) | |
| Missing | 3 (0.1%) | 1 (0.0%) | |
| **Exposures, n (%)** | | | |
| Severe isolation ( < 1) | 437 (7.8%) | 608 (6.1%) | <0.001 |
| Moderate-to-severe isolation (< 2) | 1660 (29.8%) | 2446 (24.6%) | <0.001 |
| Loneliness | 1030 (18.5%) | 1612 (16.2%) | <0.001 |

Source: Health and Retirement Study (HRS) 2016 respondents eligible for the Venous Blood Study (VBS). Values are presented as mean (SD) for continuous variables and n (%) for categorical variables. Participants were classified according to whether they completed the 2016 VBS, conditional on eligibility (i.e., participating in the 2016 HRS Core, dwelling in the community). P values were obtained from t tests for continuous variables and chi-square tests for categorical variables. Severe isolation was defined as < 1 point and moderate-to-severe isolation as < 2 points on an 5-point scale, with lower scores indicating greater social isolation. Loneliness was assessed using a single-item measure of past-week loneliness (yes/no).

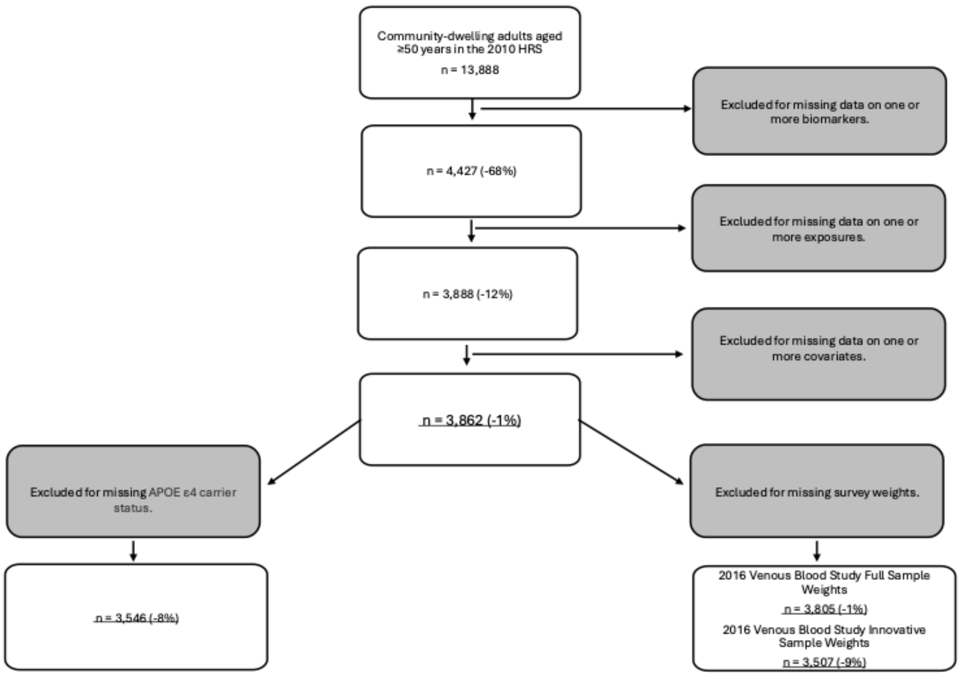

**eFigure 1. Analytic Sample Selection Process**

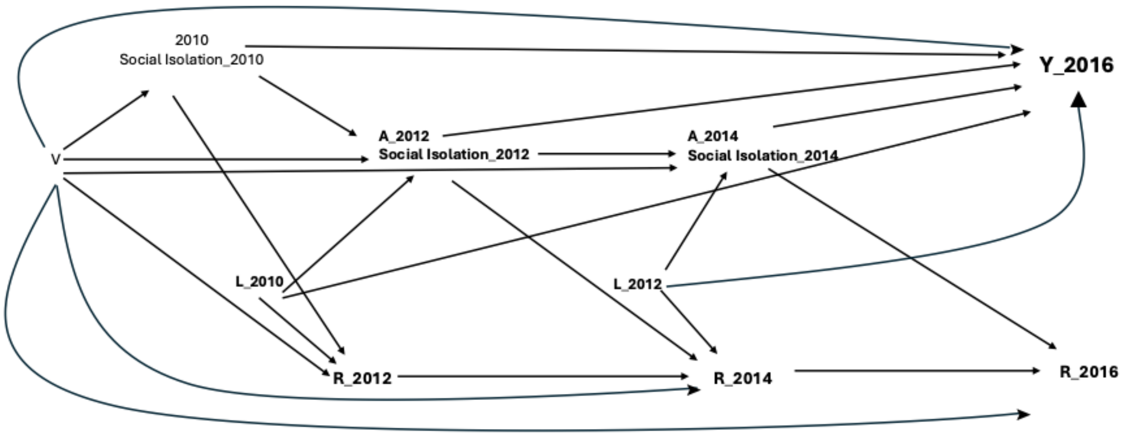

**eFigure 2. Directed acyclic graph illustrating the hypothesized relationships between social isolation/loneliness and AD/ADRD biomarkers.**

A denotes exposure (social isolation/loneliness), V denotes baseline covariates, L denotes time-varying covariates, R denotes censoring (e.g., loss to follow-up), and Y denotes the outcome. Arrows indicate assumed causal relationships.

#### eTable 2. Social Isolation Scale Components and Scoring

| Core HRS social isolation scale reported in Cenzer et al. | Modified version of Cenzer et al. |
| --- | --- |
| Marital status:<br>• not married or partnered (0 points)<br>• married or partnered (1 point) | Marital status<br>• not married or partnered (0 points)<br>married or partnered (1 point) |
| Household size:<br>• living alone (0 points)<br>• living with one person (1 point)<br>• living with two or more people (2 points) | Household size:<br>• living alone (0 points)<br>• living with one person (0.5 point)<br>living with two or more people (1 points) |
| Proximity to children:<br>• no children within 10 miles (0 points)<br>• one or more children within 10 miles (1 point) | Proximity to children:<br>• no children within 10 miles (0 points)<br>one or more children within 10 miles (1 point) |
| Frequency of religious service attendance:<br>• none (0 points)<br>• one or more times per year (1 point)<br>• two or more times per month (2 points) | Frequency of religious service attendance:<br>• none (0 points)<br>• one or more times per year (0.5 point)<br>two or more times per month (1 points) |
| Participation in volunteer activities:<br>• none (0 points)<br>• 1-50 hours per year (1 point)<br>• 51 hours or more per year (2 points) | Participation in volunteer activities:<br>• none (0 points)<br>• 1-50 hours per year (0.5 point)<br>51 hours or more per year (1 points) |
| Total Score: 0-8<br>Severe Social Isolation: ≤ 2 points<br>Moderate-to-Severe Social Isolation: ≤ 3 points | Total Score: 0-5<br>Severe Social Isolation: < 1 point<br>Moderate-to-Severe Social Isolation: < 2 points |

**eText 1. HRS Sample Weights**

The Health and Retirement Study (HRS) provides weights to account for differential probabilities of selection and participation. For analyses involving blood-based biomarkers, two sets of weights are available. The Venous Blood Study (VBS) full-sample weights account for differential participation in the 2016 VBS, whereas the VBS Innovation Sample weights additionally account for selection into the Innovation Sample from which blood-based AD/ADRD biomarkers were assayed.

As a sensitivity analysis, we incorporated these HRS weights to evaluate the robustness of our findings to differential selection into the VBS and Innovation Sample. We conducted separate analyses using the VBS full-sample weights and the VBS Innovation Sample weights.

To facilitate integration with the inverse probability weights described in eText 2, weights were standardized by dividing each weight by its mean. The standardized weights were then multiplied by the trimmed inverse probability treatment and retention weights (IPTW × IPCW). The resulting combined weights were truncated at the 1st and 99th percentiles.
Weights were not available for all participants in the analytic sample because of nonparticipation in the VBS or invalid venous blood biomarker measures. Consequently, these sensitivity analyses were restricted to participants with nonmissing weights (n = 3805 for analyses using the VBS full-sample weights and n = 3507 for analyses using the VBS Innovation Sample weights). All weighted sensitivity analyses were conducted using z-standardized log-transformed biomarker outcomes.

**eText 2. Construction of Inverse Probability Weights**

**1. Inverse probability of treatment weights**

Stabilized inverse probability of treatment weights (IPTW) were constructed separately for each exposure definition, including social isolation defined using cutoffs of <1 and <2 points and loneliness. The covariates included in each weighting model are summarized in eTable 4.

Our primary model of social isolation takes the following form:

$$Y_{2016} = \beta_0 + \beta_1\, Social\ Isolation_{2012} + \beta_2\, Social\ Isolation_{2014} + \beta_3 V + \varepsilon_i$$

$Y_{2016}$ reflects blood-based AD/ADRD biomarker values measured in the 2016 Venous Blood Innovation Study. Our exposures of interest are social isolation measured at two and four years before outcome ascertainment. $V$ captures a limited set of baseline covariates measured in 2010, including age, sex, race and ethnicity, and educational attainment.

To address time-varying confounding, we estimated pooled logistic regression models for the probability of social isolation at each wave, conditional on prior exposure history, time-varying covariates lagged to one wave prior, and baseline covariates:

$$\text{logit}[\Pr(Social\ Isolation_t = 1\ |Social\ Isolation_{t-1},\ L_{t-1}, V)] = \beta_0 + \beta_1 Social\ Isolation_{t-1} + \beta_2 L_{t-1} + \beta_3 V + \beta_4 Study\ Visit_t + \varepsilon_i$$

Stabilized numerator models included only baseline covariates:

$$\text{logit}\ [Social\ Isolation_t = 1\ |\ V)] = \alpha_0 + \alpha_2 V + \varepsilon_i$$

We used the predicted probabilities from these models to calculate stabilized weights as:

$$\widehat{sw}_t^i = \frac{\widehat{Pr}_{num}\ (Social\ Isolation_t^i = social\ isolation_t)}{\widehat{Pr}_{den}\ (Social\ Isolation_t^i = social\ isolation_t)}$$

For each respondent, weights were calculated at each wave based on observed exposure status. The final stabilized weight was the product of wave-specific weights i.e., $\widehat{SW}^{i} = \widehat{sw}_{2012}^{i} * \widehat{sw}_{2014}^{i}$.

These same procedures were followed for models of loneliness, applied to the final outcome model:

$$Y_{2016} = \beta_0 + \beta_1 Loneliness_{2012} + \beta_2 \, Loneliness_{2014} + \beta_3 V + \varepsilon_i$$

#### 2. Inverse probability of retention weights

We additionally created a set of inverse probability of retention weights (IPRW) to account for selective retention following 2010 and through outcome assessment in 2016. Stabilized weights were calculated to account for retention at 2012, 2014, and 2016 waves, conditional on being observed at the prior wave. The covariates included in each weighting model are summarized in eTable 4.

We estimated pooled logistic regression models for retention at each wave, where $R_t$ = 1 denotes retention at wave t, $A_{t-1}$ denotes prior exposure history, $X_{t-1}$ denotes a vector of time-varying covariates hypothesized to contribute to both the probability of retention in the study and biomarker values, and $V$ denotes a limited set of baseline (time-invariant) covariates, including age, sex, race and ethnicity, and educational attainment.

Denominator models were calculated as follows:

$$logit[\Pr( R_t = 1 \mid R_{t-1} = 1, A_{t-1}, X_{t-1}, V ) \mid ]$$
$$= \beta_0 + \beta_1 A_{t-1} + \beta_2 X_{t-1} + \beta_3 V + B_4 \, StudyVisit_t + \varepsilon_i$$

Stabilized numerator models included only baseline covariates and study visit:

$$logit[\Pr(R_t = 1 \mid R_{t-1} = 1,\ A_{t-1}, V) \mid ] = \alpha_0 + \ \alpha_1 V + \ \alpha_2 StudyVisit_t + \ \varepsilon_i$$

Using the predicted probabilities from the models above, inverse probability of retention weights were calculated at each wave as:

$$\widehat{Rw}_t^i = \frac{\widehat{Pr}_{num}\left(R_t^i = 1\right)}{\widehat{Pr}_{den}\left(R_t^i = 1\right)}$$

The final retention weight was calculated as the product of wave-specific retention weights:

$$\widehat{RW}^i = \widehat{Rw}_{2012}^i * \widehat{Rw}_{2014}^i * \widehat{Rw}_{2016}^i$$

**3. Final weight**

The final weight was calculated as the product of the stabilized treatment and retention weights:

$$\widehat{W}^i = \widehat{SW}^i * \widehat{RW}^i$$

The combined weights were truncated at the 1st and 99th percentiles to reduce the influence of extreme observations.

**eTable 3. Covariates included in inverse probability weighting models (denominator and numerator specifications)**

| | IPTW Denominator | IPTW/IPRW Numerator | IPTW Extended Denominator | IPTW Extended Denominator | IPRW Denominator | IPRW Extended Denominator |
|---|---|---|---|---|---|---|
| Age | ✓ | ✓ | ✓ | ✓ | ✓ | ✓ |
| Sex | ✓ | ✓ | ✓ | ✓ | ✓ | ✓ |
| Race/Ethnicity | ✓ | ✓ | ✓ | ✓ | ✓ | ✓ |
| Educational attainment | ✓ | ✓ | ✓ | ✓ | ✓ | ✓ |
| Mother's educational attainment | ✓ | | ✓ | ✓ | ✓ | ✓ |
| Father's educational attainment | ✓ | | ✓ | ✓ | ✓ | ✓ |
| Marital status | ‡ | | ‡ | ‡ | ✓ | ✓ |
| Spouse's educational attainment | ✓ | | ✓ | ✓ | ✓ | ✓ |
| Chronic health conditions | ✓ | | ✓ | ✓ | ✓ | ✓ |
| Self-rated hearing | ✓ | | ✓ | ✓ | ✓ | ✓ |
| Self-rated vision | ✓ | | ✓ | ✓ | ✓ | ✓ |
| Cognitive status | ✓ | | ✓ | ✓ | ✓ | ✓ |
| Current smoking | | | ✓ | | ✓ | ✓ |
| Current alcohol use | | | ✓ | | ✓ | ✓ |
| Any weekly physical activity | | | ✓ | | ✓ | ✓ |
| Elevated depression | | | ✓ | | ✓ | ✓ |
| Lagged Social Isolation | ✓ | | ✓ | ✓ | ✓ | ✓ |
| Lagged Loneliness | ✓ | | ✓ | ✓ | ✓ | ✓ |
| APOE ε4 carrier status | | | | ✓ | | ✓ |

Note: ‡Included only in loneliness models.

### eTable 4. Definitions and Measurement of Covariates

| Baseline Covariates | |
|---|---|
| Age | Continuous |
| Sex | women / men |
| Race/Ethnicity | Non-Hispanic White / Non-Hispanic Black / Hispanic / Other |
| Educational attainment | years |
| Mother's educational attainment | at or below the mean / above the mean / missing |
| Father's educational attainment | at or below the mean / above the mean / missing |
| Marital status | married or partnered / separated or divorced / widowed / never married |
| Spouse educational attainment | at or below the mean / above the mean / missing |
| **Time-varying covariates** | |
| Chronic health conditions | 0 conditions / 1–2 conditions / ≥3 conditions (count of hypertension, diabetes, cancer, lung disease, heart disease, stroke, psychiatric condition, arthritis) |
| Self-rated hearing | Excellent or very good / good / fair or poor / missing |
| Self-rated vision (worst of near and distance) | Excellent or very good / good / fair or poor / missing |
| Cognitive status | dementia / CIND / normal (Langa-Weir classification) |
| Current smoking | yes / no / missing |
| Current alcohol use | yes / no / missing |
| Any weekly physical activity | yes / no / missing |
| Elevated depression | yes / no / missing (>4 depressive symptoms, excluding the loneliness item) |

**eText 3. Additional Adjustment for Health Behaviors and Depressive Symptoms**

As a sensitivity analysis, we expanded the set of covariates included in the denominator of the inverse probability of treatment weight (IPTW) models to incorporate time-varying measures of health behaviors and depressive symptoms. Specifically, denominator models additionally adjusted for lagged measures of smoking status, alcohol consumption, physical activity, and depressive symptoms, while numerator models remained unchanged. All other aspects of the weighting and modeling strategy were identical to those used in the primary analyses. Consistent with the primary analyses, these models were estimated without incorporation of HRS weights. All sensitivity analyses were conducted using z-standardized log-transformed biomarker outcomes.

**eText 4. Additional Adjustment for APOE ε4 Carrier Status**

As a sensitivity analysis, we expanded the set of covariates included in the denominator of the inverse probability of treatment weight (IPTW) and inverse probability of retention weight (IPCW) models to incorporate APOE ε4 carrier status. These analyses were restricted to participants with non-missing APOE ε4 data (n = 3546) and were estimated without incorporation of HRS weights, consistent with the primary analyses. All other aspects of the weighting and modeling strategy remained unchanged. Biomarker outcomes were z-standardized following log-transformation.

**eTable 5. Baseline (2010) descriptive characteristics of HRS respondents 50+ participating in biennial waves from 2010 to 2016 and with blood-based AD/ADRD biomarker data**

| | Unweighted (n=3862) | Weighted using 2016 VBS Full Sample Weights (n=3805) | Weighted using 2016 VBS Innovative Sample Weights (n=3507) |
|---|---|---|---|
| **Baseline covariates** | | | |
| Age, mean (SD) | 63.8 (9.5) | 62.7 (9.2) | 62.7 (9.2) |
| Female, n (%) | 2,274 (58.9%) | 2,100 (55.2%) | 1,914 (54.6%) |
| Race/Ethnicity, n (%) | | | |
| Non-Hispanic White | 2,581 (66.8%) | 3,033 (79.7%) | 2,762 (78.8%) |
| Non-Hispanic Black | 638 (16.5%) | 326 (8.6%) | 330 (9.4%) |
| Hispanic | 537 (13.9%) | 336 (8.8%) | 312 (8.9%) |
| Other | 106 (2.7%) | 110 (2.9%) | 103 (2.9%) |
| Educational attainment (yrs.) | 12.9 (3.2) | 13.4 (3.0) | 13.3 (3.0) |
| Father's educational attainment, n (%) | | | |
| ≤ mean | 1,464 (37.9%) | 1,270 (33.4%) | 1,198 (34.2%) |
| > mean | 1,854 (48.0%) | 2,077 (54.6%) | 1,893 (54.0%) |
| Missing | 544 (14.1%) | 458 (12.0%) | 416 (11.9%) |
| Mother's educational attainment, n (%) | | | |
| ≤ mean | 1,372 (35.5%) | 1,125 (29.6%) | 1,066 (30.4%) |
| > mean | 2,197 (56.9%) | 2,430 (63.9%) | 2,212 (63.1%) |
| Missing | 293 (7.6%) | 251 (6.6%) | 229 (6.5%) |
| Marital status, n (%) | | | |
| Married/Partnered | 2,662 (68.9%) | 2,636 (69.3%) | 2,435 (69.4%) |
| Divorced/Separated | 530 (13.7%) | 517 (13.6%) | 482 (13.7%) |
| Widowed | 468 (12.1%) | 376 (9.9%) | 352 (10.0%) |
| Never married | 202 (5.2%) | 276 (7.3%) | 238 (6.8%) |
| Spouse's educational attainment, n (%) | | | |
| ≤ mean | 1,281 (33.2%) | 1,116 (29.3%) | 1,038 (29.6%) |
| > mean | 1,335 (34.6%) | 1,476 (38.8%) | 1,356 (38.7%) |
| Missing | 1,246 (32.3%) | 1,214 (31.9%) | 1,113 (31.7%) |
| **Time-varying covariates** | | | |
| Chronic health conditions, n (%) | 670 (17.3%) | 734 (19.3%) | 675 (19.2%) |
| 0 conditions | 2,065 (53.5%) | 2,044 (53.7%) | 1,872 (53.4%) |
| 1-2 conditions | 1,127 (29.2%) | 1,028 (27.0%) | 961 (27.4%) |
| ≥3 conditions | | | |
| Self-rated hearing | 1,851 (47.9%) | 1,933 (50.8%) | 1,760 (50.2%) |
| Excellent or very good | 1,324 (34.3%) | 1,229 (32.3%) | 1,142 (32.6%) |
| Good | 686 (17.8%) | 643 (16.9%) | 605 (17.2%) |
| Fair or Poor | 1 (0.0%) | | |
| Missing | | | |
| Self-rated vision | 1,328 (34.4%) | 1,451 (38.1%) | 1,321 (37.7%) |
| Excellent or very good | 1,699 (44.0%) | 1,597 (42.0%) | 1,476 (42.1%) |
| Good | 829 (21.5%) | 750 (19.7%) | 703 (20.1%) |
| Fair or Poor | 6 (0.2%) | 7 (0.2%) | 6 (0.2%) |
| Missing | | | |
| Cognitive status, n (%) | | | |
| Cognitively impaird, not dementia | 477 (12.4%) | 382 (10.0%) | 348 (9.9%) |
| Dementia | 66 (1.7%) | 55 (1.5%) | 53 (1.5%) |
| Current smoking, n (%) | 557 (14.4%) | 516 (13.6%) | 461 (13.2%) |
| Missing | 17 (0.4%) | 11 (0.3%) | 10 (0.3%) |
| Current Drinking, , n (%) | 2,379 (61.6%) | 2,478 (65.1%) | 2,271 (64.7%) |
| Any weekly physical activity, n (%) | 3,560 (92.2%) | 3,531 (92.8%) | 3,247 (92.6%) |
| Missing | 190 (4.9%) | 179 (4.7%) | 170 (4.8%) |
| Elevated depression, n (%) | 292 (7.6%) | 264 (6.9%) | 245 (7.0%) |
| Source: Health and Retirement Study (HRS), 2010-2016. | | | |

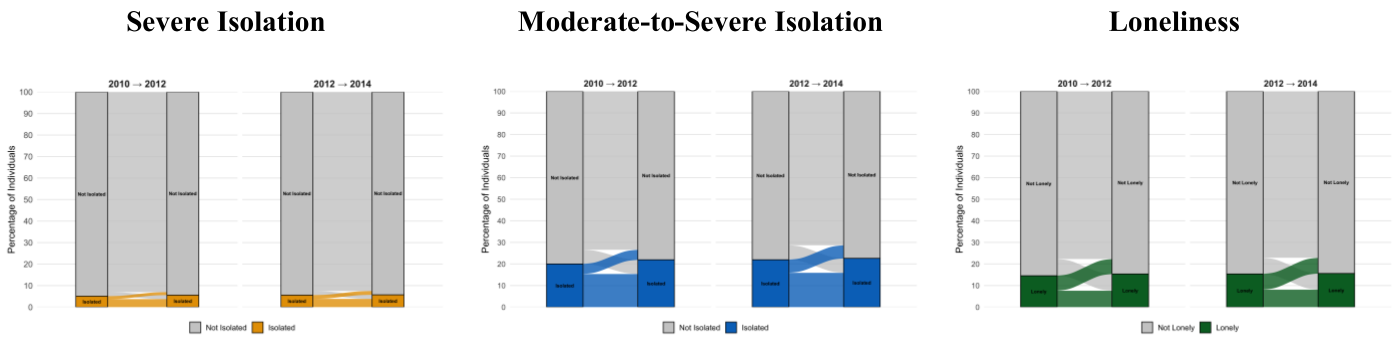

**eFigure 3. Transitions in social isolation and loneliness across study waves.**
Data are from the U.S. Health and Retirement Study (HRS), 2010–2016; (n=3862). Severe isolation was defined as <1 point and moderate-to-severe isolation as <2 points on an 5-point scale; loneliness was assessed separately. Panels show transitions between consecutive waves (2010–2012 to 2012–2014 and 2012–2014 to 2014–2016).

**eTable 6. Prevalence of four-category multi-wave isolation and loneliness exposure variables in the U.S. Health and Retirement Study.**

| | Unweighted (n=3862) | Weighted using 2016 VBS Full Sample Weights (n=3805) | Weighted Using 2016 VBS Innovative Sample Weights (n=3507) |
|---|---|---|---|
| Severe Isolation | | | |
| Persistent | 148 (3.8%) | 168 (4.4%) | 142 (4.0%) |
| Resolving | 68 (1.8%) | 78 (2.1%) | 73 (2.1%) |
| New-onset | 73 (1.9%) | 80 (2.1%) | 71 (2.0%) |
| No exposure | 3,573 (92.5%) | 3,479 (91.4%) | 3,221 (91.8%) |
| Moderate-to-Severe Isolation | | | |
| Persistent | 614 (15.9%) | 697 (18.3%) | 628 (17.9%) |
| Resolving | 234 (6.1%) | 238 (6.3%) | 218 (6.2%) |
| New-onset | 260 (6.7%) | 237 (6.2%) | 216 (6.2%) |
| No exposure | 2,754 (71.3%) | 2,633 (69.2%) | 2,445 (69.7%) |
| Loneliness | | | |
| Persistent | 313 (8.1%) | 282 (7.4%) | 250 (7.1%) |
| Resolving | 281 (7.3%) | 287 (7.5%) | 257 (7.3%) |
| New-onset | 290 (7.5%) | 249 (6.5%) | 237 (6.7%) |
| No exposure | 2,978 (77.1%) | 2,987 (78.5%) | 2,764 (78.8%) |

Source: Health and Retirement Study (HRS), 2010-2016. Severe isolation was defined as < 1 point and moderate-to-severe isolation as <2 points on an 5-point scale, with lower scores indicating more social isolation; past-week loneliness via a single-item (yes/no). Four-category multi-wave variables for severe isolation (<1 point), moderate-to-severe isolation (<2 points), and loneliness were constructed using assessments from 2012 and 2014 to classify participants as having no exposure at either wave (reference), exposure at both waves (persistent), exposure in 2012 but not 2014 (resolving), or exposure in 2014 both not 2012 (new-onset).

#### eTable 7. Distribution of AD/ADRD biomarker outcomes measured in 2016

| | Unweighted (n=3862) | Weighted using 2016 VBS Full Sample Weights (n=3805) | Weighted using 2016 VBS Innovative Sample Weights (n=3507) |
|---|---|---|---|
| **Biomarker, median (IQR)** | | | |
| **p-tau181** | 1.6 (1.1-2.5) | 1.5 (1.0-2.3) | 1.5 (1.0-2.4) |
| **Aβ42/40 ratio** | 0.1 (0.1-0.1) | 0.1 (0.1 - 0.7) | 0.1 (0.1 - 0.7) |
| **NfL** | 18.0 (11.9-29.2) | 16.9 (11.8 - 26.6) | 17.1 (11.8 - 26.8) |
| **GFAP** | 87.7 (56.7-134.5) | 81.9 (54.0 - 123.8) | 82.5 (54.3 - 124.8) |
| Source: Health and Retirement Study (HRS), 2010-2016. | | | |

**eTable 8. Numerical Estimates Corresponding to Figure 1: Associations between four-category multi-wave isolation and loneliness exposure variables and AD/ADRD biomarkers in the U.S. Health and Retirement Study.**

| | **p-tau181** | **Aβ42/40 ratio** | **NfL** | **GFAP** |
|---|---|---|---|---|
| **Severe Isolation** | | | | |
| **Persistent** | -0.00 | 0.07 | 0.31* | -0.31** |
| | (-0.45 - 0.45) | (-0.26 - 0.40) | (-0.04 - 0.66) | (-0.59 - -0.04) |
| **Resolving** | 0.22 | 0.05 | 0.15 | 0.07 |
| | (-0.08 - 0.52) | (-0.25 - 0.35) | (-0.07 - 0.38) | (-0.15 - 0.30) |
| **New-onset** | -0.05 | -0.25** | 0.13 | -0.05 |
| | (-0.27 - 0.18) | (-0.45 - -0.05) | (-0.08 - 0.35) | (-0.24 - 0.15) |
| **Moderate-to-Severe Isolation** | | | | |
| **Persistent** | -0.04 | 0.01 | 0.10 | -0.09 |
| | (-0.15 - 0.08) | (-0.12 - 0.14) | (-0.03 - 0.22) | (-0.21 - 0.03) |
| **Resolving** | 0.11* | -0.01 | 0.20*** | 0.07 |
| | (-0.02 - 0.25) | (-0.16 - 0.14) | (0.06 - 0.34) | (-0.06 - 0.19) |
| **New-onset** | 0.02 | -0.11 | 0.14** | -0.03 |
| | (-0.11 - 0.15) | (-0.24 - 0.03) | (0.01 - 0.26) | (-0.14 - 0.07) |
| **Loneliness** | | | | |
| **Persistent** | -0.01 | -0.04 | 0.11 | 0.16*** |
| | (-0.15 - 0.12) | (-0.19 - 0.10) | (-0.03 - 0.26) | (0.04 - 0.29) |
| **Resolving** | 0.02 | -0.05 | 0.10 | -0.04 |
| | (-0.09 - 0.14) | (-0.18 - 0.09) | (-0.03 - 0.22) | (-0.16 - 0.07) |
| **New-onset** | -0.01 | 0.04 | 0.07 | 0.08 |
| | (-0.13 - 0.11) | (-0.09 - 0.17) | (-0.02 - 0.17) | (-0.02 - 0.19) |

Source: Health and Retirement Study (HRS), 2010–2016. (n=3862). Severe isolation was defined as <1 point and moderate-to-severe isolation as <2 points on an 5-point scale, with lower scores indicating more social isolation; past-week loneliness via a single-item (yes/no). Four-category multi-wave variables for severe isolation (<1 point), moderate-to-severe isolation (<2 points), and loneliness were constructed using assessments from 2012 and 2014 to classify participants as having no exposure at either wave (reference), exposure at both waves (persistent), exposure in 2012 but not 2014 (resolving), or exposure in 2014 both not 2012 (new-onset). Estimates are based on linear regression models with stabilized inverse probability of treatment and retention weights and robust standard errors. Models adjust for baseline covariates (age, sex, race/ethnicity, and educational attainment). Biomarker outcomes are natural log-transformed. Estimates and 95% confidence intervals, shown in parentheses, represent percentage differences obtained by exponentiating model coefficients. *** p < 0.01, ** p < 0.05, * p < 0.1.

**eTable 9. Associations between four-category multi-wave isolation and loneliness exposure variablesand AD/ADRD biomarkers in the U.S. Health and Retirement Study, with AD/ADRD biomarkers expressed as percentage differences based on log-transformed outcomes**

| | p-tau181 | Aβ42/40 ratio | NfL | GFAP |
|---|---|---|---|---|
| **Severe Isolation** | | | | |
| **Persistent** | -0.13 | 1.81 | 24.27* | -18.28** |
| | (-27.59 – 37.74) | (-6.28 – 10.60) | (-2.83 – 58.93) | (-31.65 – -2.31) |
| **Resolving** | 16.94 | 1.21 | 11.2 | 4.98 |
| | (-5.66 – 44.97) | (-6.12 – 9.12) | (-5.07 – 30.25) | (-9.43 – 21.67) |
| **New-onset** | -3.21 | -6.00** | 9.79 | -3.08 |
| | (-17.57 – 13.64) | (-10.59 – -1.18) | (-5.63 – 27.73) | (-14.66 – 10.06) |
| **Moderate-to-Severe Isolation** | | | | |
| **Persistent** | -2.53 | 0.23 | 7.11 | -5.75 |
| | (-9.97 – 5.53) | (-2.99 – 3.56) | (-1.82 – 16.86) | (-12.73 – 1.78) |
| **Resolving** | 8.45* | -0.25 | 14.88*** | 4.48 |
| | (-1.34 – 19.20) | (-3.85 – 3.49) | (3.99 – 26.91) | (-3.67 – 13.31) |
| **New-onset** | 1.43 | -2.68 | 10.2** | -2.25 |
| | (-7.30 – 10.98) | (-5.95 – 0.71) | (0.96 – 20.29) | (-8.93 – 4.93) |
| **Loneliness** | | | | |
| **Persistent** | -0.95 | -1.11 | 8.1 | 11.18*** |
| | (-10.01 – 9.01) | (-4.57 – 2.46) | (-2.36 – 19.69) | (2.58 – 20.50) |
| **Resolving** | 1.63 | -1.13 | 6.88 | -2.77 |
| | (-6.40 – 10.35) | (-4.43 – 2.29) | (-1.88 – 16.42) | (-9.61 – 4.60) |
| **New-onset** | -0.76 | 0.99 | 5.26 | 5.59 |
| | (-8.82 – 8.00) | (-2.19 – 4.28) | (-1.68 – 12.69) | (-1.19 – 12.83) |

Source: Health and Retirement Study (HRS), 2010–2016. (n=3862). Severe isolation was defined as < 1 point and moderate-to-severe isolation as < 2 points on an 5-point scale, with lower scores indicating more social isolation; past-week loneliness via a single-item (yes/no). Four-category multi-wave variables for severe isolation (<1 point), moderate-to-severe isolation (<2 points), and loneliness were constructed using assessments from 2012 and 2014 to classify participants as having no exposure at either wave (reference), exposure at both waves (persistent), exposure in 2012 but not 2014 (resolving), or exposure in 2014 both not 2012 (new-onset). Estimates are based on linear regression models with stabilized inverse probability of treatment and retention weights and robust standard errors. Models adjust for baseline covariates (age, sex, race/ethnicity, and educational attainment). Biomarker outcomes are natural log-transformed. Estimates and 95% confidence intervals, shown in parentheses, represent percentage differences obtained by exponentiating model coefficients. *** p < 0.01, ** p < 0.05, * p < 0.1.

**eTable 10. Associations between four-category multi-wave isolation and loneliness exposure variables and AD/ADRD biomarkers in the U.S. Health and Retirement Study, with AD/ADRD biomarkers expressed as z-standardized units**

| | p-tau181 | Aβ42/40 ratio | NfL | GFAP |
|---|---|---|---|---|
| **Severe Isolation** | | | | |
| **Persistent** | -0.05 | 0.02 | 0.44 | -0.25** |
| | (-0.29 - 0.18) | (-0.23 - 0.26) | (-0.13 - 1.01) | (-0.45 - -0.04) |
| **Resolving** | 0.25 | 0.04 | 0.08 | 0.12 |
| | (-0.06 - 0.55) | (-0.19 - 0.27) | (-0.11 - 0.27) | (-0.25 - 0.49) |
| **New-onset** | -0.04 | -0.22*** | 0.13 | -0.06 |
| | (-0.23 - 0.14) | (-0.36 - -0.07) | (-0.14 - 0.39) | (-0.28 - 0.16) |
| **Moderate-to-Severe Isolation** | | | | |
| **Persistent** | -0.09** | -0.00 | 0.10 | -0.05 |
| | (-0.18 - -0.00) | (-0.11 - 0.11) | (-0.04 - 0.24) | (-0.16 - 0.07) |
| **Resolving** | 0.03 | -0.02 | 0.19* | 0.12 |
| | (-0.10 - 0.16) | (-0.14 - 0.09) | (-0.03 - 0.40) | (-0.07 - 0.31) |
| **New-onset** | -0.08* | -0.09* | 0.19 | -0.06 |
| | (-0.17 - 0.01) | (-0.19 - 0.01) | (-0.09 - 0.47) | (-0.17 - 0.05) |
| **Loneliness** | | | | |
| **Persistent** | 0.01 | -0.05 | 0.18 | 0.18** |
| | (-0.12 - 0.15) | (-0.17 - 0.07) | (-0.18 - 0.54) | (0.03 - 0.33) |
| **Resolving** | 0.01 | -0.04 | 0.06 | -0.01 |
| | (-0.16 - 0.18) | (-0.14 - 0.06) | (-0.04 - 0.17) | (-0.11 - 0.09) |
| **New-onset** | -0.03 | 0.04 | 0.04 | 0.05 |
| | (-0.12 - 0.07) | (-0.09 - 0.16) | (-0.06 - 0.14) | (-0.06 - 0.15) |

Source: Health and Retirement Study (HRS), 2010–2016. (n=3862). Severe isolation was defined as < 1 point and moderate-to-severe isolation as < 2 points on an 5-point scale, with lower scores indicating more social isolation; past-week loneliness via a single-item (yes/no). Four-category multi-wave variables for severe isolation (<1 point), moderate-to-severe isolation (<2 points), and loneliness were constructed using assessments from 2012 and 2014 to classify participants as having no exposure at either wave (reference), exposure at both waves (persistent), exposure in 2012 but not 2014 (resolving), or exposure in 2014 both not 2012 (new-onset). Estimates are based on linear regression models with stabilized inverse probability of treatment and retention weights and robust standard errors. Models adjust for baseline covariates (age, sex, race/ethnicity, and educational attainment). Biomarker outcomes are z-standardized, and estimates represent standardized differences in biomarker levels (in standard deviation units).95% confidence intervals are shown in parenthesis. *** p < 0.01, ** p < 0.05, * p < 0.1.

#### eTable 11. Numerical Estimates Corresponding to Figure 2: Associations between four-category multi-wave isolation and loneliness exposure variables and AD/ADRD biomarkers, Stratified by Sex, in the U.S. Health and Retirement Study

| | Women | | | | Men | | | |
|---|---|---|---|---|---|---|---|---|
| | p-tau181 | Aβ42/40 ratio | NfL | GFAP | p-tau181 | Aβ42/40 ratio | NfL | GFAP |
| **Severe Isolation** | | | | | | | | |
| **Persistent** | -0.39 | -0.15 | 0.39 | -0.34** | 0.41*** | 0.33 | 0.21 | -0.26 |
| | (-1.04 - 0.26) | (-0.60 - 0.31) | (-0.11 - 0.88) | (-0.65 - -0.04) | (0.12 - 0.69) | (-0.10 - 0.75) | (-0.27 - 0.70) | (-0.72 - 0.20) |
| **Resolving** | 0.39** | -0.03 | 0.19 | 0.09 | -0.09 | 0.19 | 0.08 | 0.05 |
| | (0.02 - 0.75) | (-0.43 - 0.37) | (-0.10 - 0.48) | (-0.21 - 0.39) | (-0.57 - 0.39) | (-0.25 - 0.64) | (-0.26 - 0.43) | (-0.29 - 0.38) |
| **New-onset** | -0.03 | -0.20 | 0.14 | -0.18 | -0.06 | -0.31** | 0.12 | 0.11 |
| | (-0.34 - 0.28) | (-0.48 - 0.08) | (-0.18 - 0.47) | (-0.43 - 0.08) | (-0.39 - 0.27) | (-0.58 - -0.04) | (-0.15 - 0.39) | (-0.18 - 0.40) |
| **Moderate-to-Severe Isolation** | | | | | | | | |
| **Persistent** | -0.05 | -0.06 | 0.11 | -0.09 | -0.01 | 0.14 | 0.07 | -0.10 |
| | (-0.20 - 0.10) | (-0.22 - 0.10) | (-0.05 - 0.27) | (-0.25 - 0.08) | (-0.16 - 0.14) | (-0.09 - 0.37) | (-0.12 - 0.27) | (-0.26 - 0.05) |
| **Resolving** | 0.11 | -0.04 | 0.17* | 0.02 | 0.12 | 0.04 | 0.26** | 0.16 |
| | (-0.04 - 0.26) | (-0.21 - 0.13) | (-0.00 - 0.34) | (-0.12 - 0.15) | (-0.14 - 0.38) | (-0.24 - 0.33) | (0.01 - 0.51) | (-0.08 - 0.41) |
| **New-onset** | 0.03 | -0.14 | 0.12 | -0.04 | -0.01 | -0.05 | 0.15 | -0.02 |
| | (-0.14 - 0.20) | (-0.33 - 0.05) | (-0.04 - 0.29) | (-0.17 - 0.08) | (-0.19 - 0.17) | (-0.23 - 0.13) | (-0.05 - 0.36) | (-0.21 - 0.18) |
| **Loneliness** | | | | | | | | |
| **Persistent** | 0.01 | -0.07 | 0.12 | 0.20*** | -0.03 | 0.04 | 0.08 | 0.06 |
| | (-0.15 - 0.17) | (-0.25 - 0.10) | (-0.03 - 0.26) | (0.05 - 0.34) | (-0.28 - 0.21) | (-0.17 - 0.24) | (-0.31 - 0.47) | (-0.17 - 0.29) |
| **Resolving** | 0.11 | -0.04 | 0.14* | 0.01 | -0.14* | -0.06 | -0.04 | -0.17 |
| | (-0.04 - 0.26) | (-0.20 - 0.12) | (-0.01 - 0.29) | (-0.12 - 0.14) | (-0.31 - 0.02) | (-0.32 - 0.20) | (-0.26 - 0.18) | (-0.39 - 0.04) |
| **New-onset** | 0.01 | 0.10 | -0.01 | 0.01 | -0.05 | -0.07 | 0.26*** | 0.25*** |
| | (-0.14 - 0.17) | (-0.07 - 0.26) | (-0.12 - 0.10) | (-0.11 - 0.13) | (-0.23 - 0.13) | (-0.25 - 0.12) | (0.07 - 0.45) | (0.07 - 0.44) |

Source: Health and Retirement Study (HRS), 2010–2016; (Women: N = 2,274; Men: N = 1,588). Severe isolation was defined as < 1 point and moderate-to-severe isolation as < 2 points on an 5-point scale, with lower scores indicating more social isolation; past-week loneliness via a single-item (yes/no). Four-category multi-wave variables for severe isolation (<1 point), moderate-to-severe isolation (<2 points), and loneliness were constructed using assessments from 2012 and 2014 to classify participants as having no exposure at either wave (reference), exposure at both waves (persistent), exposure in 2012 but not 2014 (resolving), or exposure in 2014 both not 2012 (new-onset). Estimates are shown stratified by sex. Estimates are based on linear regression models with stabilized inverse probability of treatment and retention weights and robust standard errors. Models adjust for baseline covariates (age, sex, race/ethnicity, and educational attainment). P values for interactions: severe isolation: persistent (p-tau181 = .02, Aβ42/40 = .16, NfL = .60, GFAP = .79), resolving (p-tau181 = .13, Aβ42/40 = .48, NfL = .62, GFAP = .84), and new-onset (p-tau181 = .89, Aβ42/40 = .57, NfL = .91, GFAP = .14); moderate-to-severe isolation: persistent (p-tau181 = .55, Aβ42/40 = .23, NfL = .65, GFAP = .76), resolving (p-tau181 = .89, Aβ42/40 = .69, NfL = .63, GFAP = .39), and new-onset (p-tau181 = .82, Aβ42/40 = .61, NfL = .85, GFAP = .98); loneliness: persistent (p-tau181 = .99, Aβ42/40 = .54, NfL = .85, GFAP = .25), resolving (p-tau181 = .05, Aβ42/40 = .80, NfL = .19, GFAP = .10), and new-onset (p-tau181 = .73, Aβ42/40 = .17, NfL = .02, GFAP = .03).
*** p<0.01, ** p<0.05, * p<0.1.

**eTable 12. Numerical Estimates Corresponding to Figure 3: Associations between four-category multi-wave isolation and loneliness exposure variables and AD/ADRD biomarkers, Stratified by APOE ε4 carrier status, in the U.S. Health and Retirement Study.**

| | APOE ε4 Carrier | | | | APOE ε4 Non-Carrier | | | |
|---|---|---|---|---|---|---|---|---|
| | p-tau181 | Aβ42/40 ratio | NfL | GFAP | p-tau181 | Aβ42/40 ratio | NfL | GFAP |
| **Severe Isolation** | | | | | | | | |
| **Persistent** | -0.11 | 0.61** | 0.14* | 0.03 | -0.08 | -0.16 | 0.48* | -0.44*** |
| | (-1.09 - 0.88) | (0.12 - 1.10) | (-0.01 - 0.30) | (-0.31 - 0.37) | (-0.68 - 0.53) | (-0.56 - 0.23) | (-0.04 - 0.99) | (-0.76 - -0.12) |
| **Resolving** | -0.43*** | 0.48* | -0.20 | 0.00 | 0.25 | -0.18 | 0.20 | 0.03 |
| | (-0.75 - -0.11) | (-0.06 - 1.02) | (-0.71 - 0.31) | (-0.51 - 0.52) | (-0.09 - 0.59) | (-0.52 - 0.15) | (-0.06 - 0.45) | (-0.21 - 0.26) |
| **New-Onset** | 0.16 | 0.05 | 0.34* | 0.11 | -0.05 | -0.31** | 0.11 | -0.10 |
| | (-0.35 - 0.67) | (-0.30 - 0.40) | (-0.03 - 0.71) | (-0.25 - 0.46) | (-0.31 - 0.22) | (-0.56 - -0.06) | (-0.15 - 0.37) | (-0.32 - 0.12) |
| **Moderate-to-Severe Isolation** | | | | | | | | |
| **Persistent** | -0.39*** | 0.05 | -0.12 | -0.28* | 0.01 | -0.00 | 0.22** | 0.01 |
| | (-0.67 - -0.11) | (-0.13 - 0.24) | (-0.32 - 0.08) | (-0.58 - 0.02) | (-0.11 - 0.13) | (-0.18 - 0.18) | (0.05 - 0.38) | (-0.12 - 0.14) |
| **Resolving** | 0.02 | 0.20 | 0.11 | 0.10 | 0.07 | -0.11 | 0.22** | 0.08 |
| | (-0.22 - 0.27) | (-0.07 - 0.47) | (-0.15 - 0.38) | (-0.11 - 0.30) | (-0.09 - 0.23) | (-0.28 - 0.06) | (0.04 - 0.40) | (-0.08 - 0.24) |
| **New-Onset** | 0.08 | 0.10 | 0.09 | -0.07 | -0.04 | -0.15 | 0.16* | 0.01 |
| | (-0.15 - 0.31) | (-0.13 - 0.32) | (-0.10 - 0.29) | (-0.28 - 0.14) | (-0.20 - 0.13) | (-0.33 - 0.03) | (-0.01 - 0.34) | (-0.13 - 0.14) |
| **Loneliness** | | | | | | | | |
| **Persistent** | -0.17 | -0.12 | 0.18* | 0.51*** | -0.02 | -0.03 | 0.11 | 0.17** |
| | (-0.41 - 0.07) | (-0.44 - 0.20) | (-0.02 - 0.37) | (0.25 - 0.77) | (-0.20 - 0.15) | (-0.21 - 0.15) | (-0.06 - 0.27) | (0.03 - 0.32) |
| **Resolving** | 0.01 | -0.26** | 0.20 | 0.05 | -0.02 | -0.00 | 0.11 | 0.00 |
| | (-0.25 - 0.28) | (-0.51 - -0.01) | (-0.10 - 0.49) | (-0.17 - 0.26) | (-0.15 - 0.12) | (-0.16 - 0.16) | (-0.02 - 0.25) | (-0.14 - 0.14) |
| **New-Onset** | 0.05 | -0.01 | 0.04 | 0.08 | -0.07 | 0.03 | 0.09 | 0.11* |
| | (-0.22 - 0.32) | (-0.28 - 0.26) | (-0.13 - 0.21) | (-0.07 - 0.23) | (-0.21 - 0.07) | (-0.12 - 0.17) | (-0.03 - 0.21) | (-0.02 - 0.24) |

Source: Health and Retirement Study (HRS), 2010–2016; (APOE ε4 carrier: n = 903; Non-APOE ε4 carrier: n = 2,643). Severe isolation was defined as <1 point and moderate-to-severe isolation as <2 points on an 5-point scale, with lower scores indicating more social isolation; past-week loneliness via a single-item (yes/no). Four-category multi-wave variables for severe isolation (<1 point), moderate-to-severe isolation (<2 points), and loneliness were constructed using assessments from 2012 and 2014 to classify participants as having no exposure at either wave (reference), exposure at both waves (persistent), exposure in 2012 but not 2014 (resolving), or exposure in 2014 both not 2012 (new-onset). Estimates are shown stratified by APOE ε4 carrier. Estimates are based on linear regression models with stabilized inverse probability of treatment and retention weights and robust standard errors. Models adjust for baseline covariates (age, sex, race/ethnicity, and educational attainment). ). P values for interactions: severe isolation: persistent (p-tau181 = .97, Aβ42/40 = .01, NfL = .25, GFAP = .03), resolving (p-tau181 = .00, Aβ42/40 = .05, NfL = .18, GFAP = .96), and new-onset (p-tau181 = .44, Aβ42/40 = .06, NfL = .31, GFAP = .28); moderate-to-severe isolation: persistent (p-tau181 = .03, Aβ42/40 = .46, NfL = .03, GFAP = .27), resolving (p-tau181 = .84, Aβ42/40 = .05, NfL = .62, GFAP = .65), and new-onset (p-tau181 = .41, Aβ42/40 = .10, NfL = .62, GFAP = .58); loneliness: persistent (p-tau181 = .21, Aβ42/40 = .82, NfL = .82, GFAP = .04), resolving (p-tau181 = .96, Aβ42/40 = .09, NfL = .82, GFAP = .99), and new-onset (p-tau181 = .50, Aβ42/40 = .97, NfL = .49, GFAP = .84).
*** p<0.01, ** p<0.05, * p<0.1.

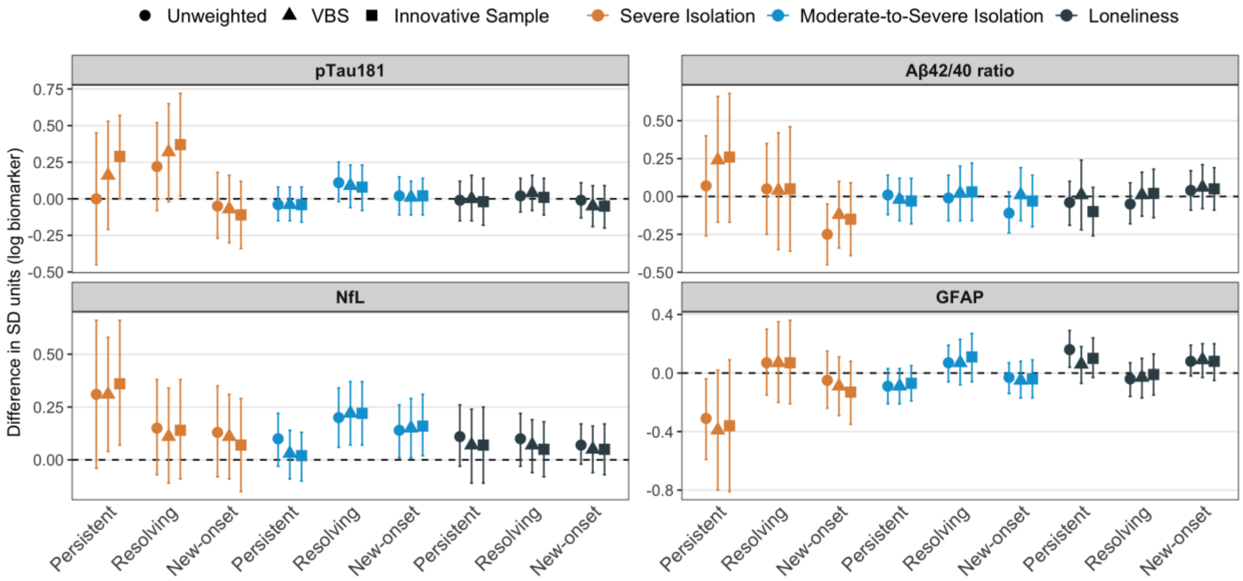

**eFigure 4. Associations between four-category multi-wave isolation and loneliness exposure variables and AD/ADRD biomarkers, using HRS Weights, in the U.S. Health and Retirement Study.**

Data are from the Health and Retirement Study (HRS), 2010-2016. Sample sizes for the primary analytic sample (unweighted), Venous Blood Study (VBS), and VBS innovative sample (B) are N=3862, N=3805, and N=3507, respectively. Points represent estimated differences in standard deviation (SD) units of log-transformed biomarker levels, and error bars indicate 95% confidence intervals. Four-category multi-wave variables for severe isolation (<1 point), moderate-to-severe isolation (<2 points), and loneliness were constructed using assessments from 2012 and 2014 to classify participants as having no exposure at either wave (reference), exposure at both waves (persistent), exposure in 2012 but not 2014 (resolving), or exposure in 2014 both not 2012 (new-onset). Estimates are based on linear regression models with stabilized inverse probability of treatment and retention weights and robust standard errors. Models adjust for baseline covariates (age, sex, race/ethnicity, and educational attainment).

**eTable 13. Numerical Estimates Corresponding to eFigure 4: Associations between four-category multi-wave isolation and loneliness exposure variables and AD/ADRD biomarkers, using HRS Weights, in the U.S. Health and Retirement Study.**

| | p-tau181 | Aβ42/40 ratio | NfL | GFAP |
|---|---|---|---|---|
| **Severe Isolation** | | | | |
| **Persistent** | | | | |
| **Unweighted** | -0.00 | 0.07 | 0.31* | -0.31** |
| | (-0.45 - 0.45) | (-0.26 - 0.40) | (-0.04 - 0.66) | (-0.59 - -0.04) |
| **2016 VBS Full Sample Weights** | 0.16 | 0.24 | 0.31** | -0.39* |
| | (-0.21 - 0.53) | (-0.17 - 0.66) | (0.04 - 0.58) | (-0.80 - 0.02) |
| **2016 VBS Innovative Sample Weights** | 0.29* | 0.26 | 0.36** | -0.36 |
| | (-0.00 - 0.57) | (-0.17 - 0.68) | (0.07 - 0.66) | (-0.81 - 0.09) |
| **Resolving** | | | | |
| **Unweighted** | 0.22 | 0.05 | 0.15 | 0.07 |
| | (-0.08 - 0.52) | (-0.25 - 0.35) | (-0.07 - 0.38) | (-0.15 - 0.30) |
| **2016 VBS Full Sample Weights** | 0.32* | 0.04 | 0.11 | 0.07 |
| | (-0.02 - 0.65) | (-0.35 - 0.42) | (-0.11 - 0.34) | (-0.20 - 0.35) |
| **2016 VBS Innovative Sample Weights** | 0.37** | 0.05 | 0.14 | 0.07 |
| | (0.02 - 0.72) | (-0.36 - 0.46) | (-0.09 - 0.38) | (-0.21 - 0.36) |
| **New-Onset** | | | | |
| **Unweighted** | -0.05 | -0.25** | 0.13 | -0.05 |
| | (-0.27 - 0.18) | (-0.45 - -0.05) | (-0.08 - 0.35) | (-0.24 - 0.15) |
| **2016 VBS Full Sample Weights** | -0.07 | -0.12 | 0.11 | -0.09 |
| | (-0.30 - 0.16) | (-0.34 - 0.10) | (-0.09 - 0.31) | (-0.29 - 0.11) |
| **2016 VBS Innovative Sample Weights** | -0.11 | -0.15 | 0.07 | -0.13 |
| | (-0.34 - 0.12) | (-0.39 - 0.09) | (-0.15 - 0.29) | (-0.35 - 0.08) |
| **Moderate-to-Severe Isolation** | | | | |
| **Persistent** | | | | |
| **Unweighted** | -0.04 | 0.01 | 0.10 | -0.09 |
| | (-0.15 - 0.08) | (-0.12 - 0.14) | (-0.03 - 0.22) | (-0.21 - 0.03) |
| **2016 VBS Full Sample Weights** | -0.04 | -0.02 | 0.03 | -0.09 |
| | (-0.15 - 0.08) | (-0.16 - 0.12) | (-0.09 - 0.14) | (-0.21 - 0.03) |
| **2016 VBS Innovative Sample Weights** | -0.04 | -0.03 | 0.02 | -0.07 |
| | (-0.16 - 0.08) | (-0.18 - 0.12) | (-0.10 - 0.13) | (-0.19 - 0.05) |
| **Resolving** | | | | |
| **Unweighted** | 0.11* | -0.01 | 0.20*** | 0.07 |
| | (-0.02 - 0.25) | (-0.16 - 0.14) | (0.06 - 0.34) | (-0.06 - 0.19) |
| **2016 VBS Full Sample Weights** | 0.09 | 0.02 | 0.22*** | 0.07 |
| | (-0.06 - 0.23) | (-0.16 - 0.20) | (0.07 - 0.37) | (-0.08 - 0.23) |
| **2016 VBS Innovative Sample Weights** | 0.08 | 0.03 | 0.22*** | 0.11 |
| | (-0.08 - 0.23) | (-0.16 - 0.22) | (0.07 - 0.37) | (-0.06 - 0.27) |
| **New-Onset** | | | | |
| **Unweighted** | 0.02 | -0.11 | 0.14** | -0.03 |
| | (-0.11 - 0.15) | (-0.24 - 0.03) | (0.01 - 0.26) | (-0.14 - 0.07) |
| **2016 VBS Full Sample Weights** | 0.01 | 0.01 | 0.15** | -0.05 |
| | (-0.11 - 0.12) | (-0.16 - 0.19) | (0.01 - 0.29) | (-0.17 - 0.08) |
| **2016 VBS Innovative Sample Weights** | 0.02 | -0.03 | 0.16** | -0.04 |
| | (-0.11 - 0.14) | (-0.20 - 0.14) | (0.02 - 0.31) | (-0.17 - 0.09) |
| **Loneliness** | | | | |
| **Persistent** | | | | |
| **Unweighted** | -0.01 | -0.04 | 0.11 | 0.16*** |
| | (-0.15 - 0.12) | (-0.19 - 0.10) | (-0.03 - 0.26) | (0.04 - 0.29) |
| **2016 VBS Full Sample Weights** | 0.00 | 0.01 | 0.07 | 0.06 |
| | (-0.15 - 0.16) | (-0.22 - 0.24) | (-0.11 - 0.24) | (-0.07 - 0.18) |
| **2016 VBS Innovative Sample Weights** | -0.02 | -0.10 | 0.07 | 0.10 |
| | (-0.18 - 0.14) | (-0.26 - 0.06) | (-0.11 - 0.25) | (-0.03 - 0.24) |
| **Resolving** | | | | |
| **Unweighted** | 0.02 | -0.05 | 0.10 | -0.04 |

| | | | | |
|---|---|---|---|---|
| | (-0.09 - 0.14) | (-0.18 - 0.09) | (-0.03 - 0.22) | (-0.16 - 0.07) |
| **2016 VBS Full Sample Weights** | 0.04 | 0.01 | 0.07 | -0.03 |
| | (-0.08 - 0.16) | (-0.13 - 0.16) | (-0.06 - 0.19) | (-0.17 - 0.10) |
| **2016 VBS Innovative Sample Weights** | 0.01 | 0.02 | 0.05 | -0.01 |
| | (-0.11 - 0.14) | (-0.14 - 0.18) | (-0.08 - 0.18) | (-0.15 - 0.13) |
| **New-Onset** | | | | |
| **Unweighted** | -0.01 | 0.04 | 0.07 | 0.08 |
| | (-0.13 - 0.11) | (-0.09 - 0.17) | (-0.02 - 0.17) | (-0.02 - 0.19) |
| **2016 VBS Full Sample Weights** | -0.05 | 0.06 | 0.05 | 0.09 |
| | (-0.19 - 0.09) | (-0.08 - 0.21) | (-0.06 - 0.16) | (-0.03 - 0.20) |
| **2016 VBS Innovative Sample Weights** | -0.05 | 0.05 | 0.05 | 0.08 |
| | (-0.20 - 0.09) | (-0.09 - 0.19) | (-0.07 - 0.17) | (-0.05 - 0.20) |

Source: Health and Retirement Study (HRS), 2010-2016. Sample sizes for the primary analytic sample (unweighted), Venous Blood Study (VBS), and VBS innovative sample (B) are n=3862, n=3805, and n=3507, respectively. Four-category multi-wave variables for severe isolation (<1 point), moderate-to-severe isolation (<2 points), and loneliness were constructed using assessments from 2012 and 2014 to classify participants as having no exposure at either wave (reference), exposure at both waves (persistent), exposure in 2012 but not 2014 (resolving), or exposure in 2014 both not 2012 (new-onset). Estimates are based on linear regression models with stabilized inverse probability of treatment and retention weights and robust standard errors. Models adjust for baseline covariates (age, sex, race/ethnicity, and educational attainment). Biomarker outcomes are natural log-transformed and subsequently z-standardized. Estimates represent standardized differences in biomarker levels. 95% confidence intervals are shown in parentheses. *** $p < 0.01$, ** $p < 0.05$, * $p < 0.1$.

**eTable 14. Associations between four-category multi-wave isolation and loneliness exposure variables and AD/ADRD biomarkers, using extended IPTW models incorporating health behaviors and depressive symptoms, in the U.S. Health and Retirement Study.**

| | p-tau181 | Aβ42/40 ratio | NfL | GFAP |
|---|---|---|---|---|
| **Severe Isolation** | | | | |
| **Persistent** | 0.04 | 0.13 | 0.36* | -0.26* |
| | (-0.40 - 0.48) | (-0.21 - 0.47) | (-0.01 - 0.73) | (-0.54 - 0.02) |
| **Resolving** | 0.23 | 0.04 | 0.16 | 0.08 |
| | (-0.07 - 0.54) | (-0.27 - 0.34) | (-0.07 - 0.38) | (-0.15 - 0.31) |
| **New-onset** | -0.01 | -0.25** | 0.12 | -0.05 |
| | (-0.24 - 0.21) | (-0.44 - -0.06) | (-0.08 - 0.33) | (-0.24 - 0.15) |
| **Moderate-to-Severe Isolation** | | | | |
| **Persistent** | -0.04 | -0.01 | 0.09 | -0.08 |
| | (-0.15 - 0.07) | (-0.13 - 0.12) | (-0.02 - 0.21) | (-0.19 - 0.03) |
| **Resolving** | 0.11* | -0.01 | 0.20*** | 0.07 |
| | (-0.02 - 0.25) | (-0.16 - 0.14) | (0.06 - 0.34) | (-0.06 - 0.19) |
| **New-onset** | 0.03 | -0.11 | 0.14** | -0.03 |
| | (-0.10 - 0.16) | (-0.24 - 0.03) | (0.02 - 0.27) | (-0.14 - 0.08) |
| **Loneliness** | | | | |
| **Persistent** | 0.03 | -0.04 | 0.15* | 0.18*** |
| | (-0.11 - 0.17) | (-0.18 - 0.09) | (-0.02 - 0.31) | (0.06 - 0.30) |
| **Resolving** | 0.02 | -0.04 | 0.09 | -0.02 |
| | (-0.10 - 0.14) | (-0.18 - 0.09) | (-0.03 - 0.21) | (-0.13 - 0.09) |
| **New-onset** | 0.01 | 0.04 | 0.07 | 0.10* |
| | (-0.11 - 0.13) | (-0.08 - 0.16) | (-0.03 - 0.16) | (-0.00 - 0.20) |

Source: Health and Retirement Study (HRS), 2010–2016. (n=3862). Severe isolation was defined as <1 point and moderate-to-severe isolation as <2 point on an 5-point scale, with lower scores indicating more social isolation; past-week loneliness via a single-item (yes/no). Four-category multi-wave variables for severe isolation (<1 point), moderate-to-severe isolation (<2 points), and loneliness were constructed using assessments from 2012 and 2014 to classify participants as having no exposure at either wave (reference), exposure at both waves (persistent), exposure in 2012 but not 2014 (resolving), or exposure in 2014 both not 2012 (new-onset). Estimates are based on linear regression models with stabilized inverse probability of treatment and retention weights and robust standard errors. Models adjust for baseline covariates (age, sex, race/ethnicity, and educational attainment). Biomarker outcomes are natural log-transformed and subsequently z-standardized. Estimates represent standardized differences in biomarker levels. 95% confidence intervals are shown in parentheses. *** p<0.01, ** p<0.05, * p<0.1.

**eTable 15. Associations between four-category multi-wave isolation and loneliness exposure variables and AD/ADRD biomarkers, using extended IPTW and IPRW models incorporating APOE ε4 carrier status, in the U.S. Health and Retirement Study.**

| | p-tau181 | Aβ42/40 ratio | NfL | GFAP |
|---|---|---|---|---|
| **Severe Isolation** | | | | |
| **Persistent** | -0.08 | 0.02 | 0.42** | -0.32** |
| | (-0.54 - 0.38) | (-0.36 - 0.39) | (0.02 - 0.81) | (-0.61 - -0.02) |
| **Resolving** | 0.10 | -0.01 | 0.11 | -0.00 |
| | (-0.18 - 0.38) | (-0.32 - 0.29) | (-0.13 - 0.34) | (-0.21 - 0.20) |
| **New-onset** | -0.01 | -0.21* | 0.16 | -0.06 |
| | (-0.26 - 0.23) | (-0.43 - 0.00) | (-0.07 - 0.38) | (-0.27 - 0.14) |
| **Moderate-to-Severe Isolation** | | | | |
| **Persistent** | -0.05 | 0.01 | 0.13* | -0.08 |
| | (-0.17 - 0.07) | (-0.12 - 0.15) | (-0.00 - 0.26) | (-0.20 - 0.05) |
| **Resolving** | 0.11 | -0.01 | 0.20** | 0.08 |
| | (-0.03 - 0.25) | (-0.16 - 0.13) | (0.05 - 0.34) | (-0.05 - 0.21) |
| **New-onset** | 0.03 | -0.08 | 0.14** | -0.06 |
| | (-0.11 - 0.16) | (-0.23 - 0.06) | (0.01 - 0.28) | (-0.17 - 0.06) |
| **Loneliness** | | | | |
| **Persistent** | -0.01 | -0.03 | 0.12* | 0.20*** |
| | (-0.16 - 0.13) | (-0.19 - 0.13) | (-0.01 - 0.26) | (0.08 - 0.33) |
| **Resolving** | 0.03 | -0.05 | 0.13** | -0.03 |
| | (-0.09 - 0.15) | (-0.19 - 0.08) | (0.01 - 0.26) | (-0.14 - 0.08) |
| **New-onset** | -0.01 | 0.04 | 0.08 | 0.06 |
| | (-0.13 - 0.12) | (-0.09 - 0.16) | (-0.02 - 0.18) | (-0.04 - 0.17) |

Source: Health and Retirement Study (HRS), 2010–2016. (n=3,546). Severe isolation was defined as <1 point and moderate-to-severe isolation as <2 points on an 5-point scale, with lower scores indicating more social isolation; past-week loneliness via a single-item (yes/no). Four-category multi-wave variables for severe isolation (<1 point), moderate-to-severe isolation (<2 points), and loneliness were constructed using assessments from 2012 and 2014 to classify participants as having no exposure at either wave (reference), exposure at both waves (persistent), exposure in 2012 but not 2014 (resolving), or exposure in 2014 both not 2012 (new-onset). Estimates are based on linear regression models with stabilized inverse probability of treatment and retention weights and robust standard errors. Models adjust for baseline covariates (age, sex, race/ethnicity, and educational attainment). Biomarker outcomes are natural log-transformed and subsequently z-standardized. Estimates represent standardized differences in biomarker levels. 95% confidence intervals are shown in parentheses. *** p<0.01, ** p<0.05, * p<0.1.

#### eTable 16. Prevalence of single-time-point social isolation and loneliness measured in 2012 and 2014

| | Unweighted (n=3862) | | Weighted (2016 VBS Full Sample Weights; n=3805) | | Weighted (2016 VBS Innovative Sample Weights;n=3507) | |
|---|---|---|---|---|---|---|
| **Single-time-point measures** | **2012** | **2014** | **2012** | **2014** | **2012** | **2014** |
| Severe isolation | 216 (5.6%) | 221 (5.7%) | 246 (6.5%) | 248 (6.5%) | 215 (6.1%) | 213 (6.1%) |
| Moderate-to-severe isolation | 848 (22.0%) | 874 (22.6%) | 935 (24.6%) | 934 (24.5%) | 846 (24.1%) | 845 (24.1%) |
| Loneliness | 594 (15.4%) | 603 (15.6%) | 569 (15.0%) | 531 (14.0%) | 507 (14.4%) | 487 (13.9%) |

Source: Health and Retirement Study (HRS), 2010-2016. Severe isolation was defined as < 1 point and moderate-to-severe isolation as <2 points on an 5-point scale, with lower scores indicating more social isolation; past-week loneliness via a single-item (yes/no).

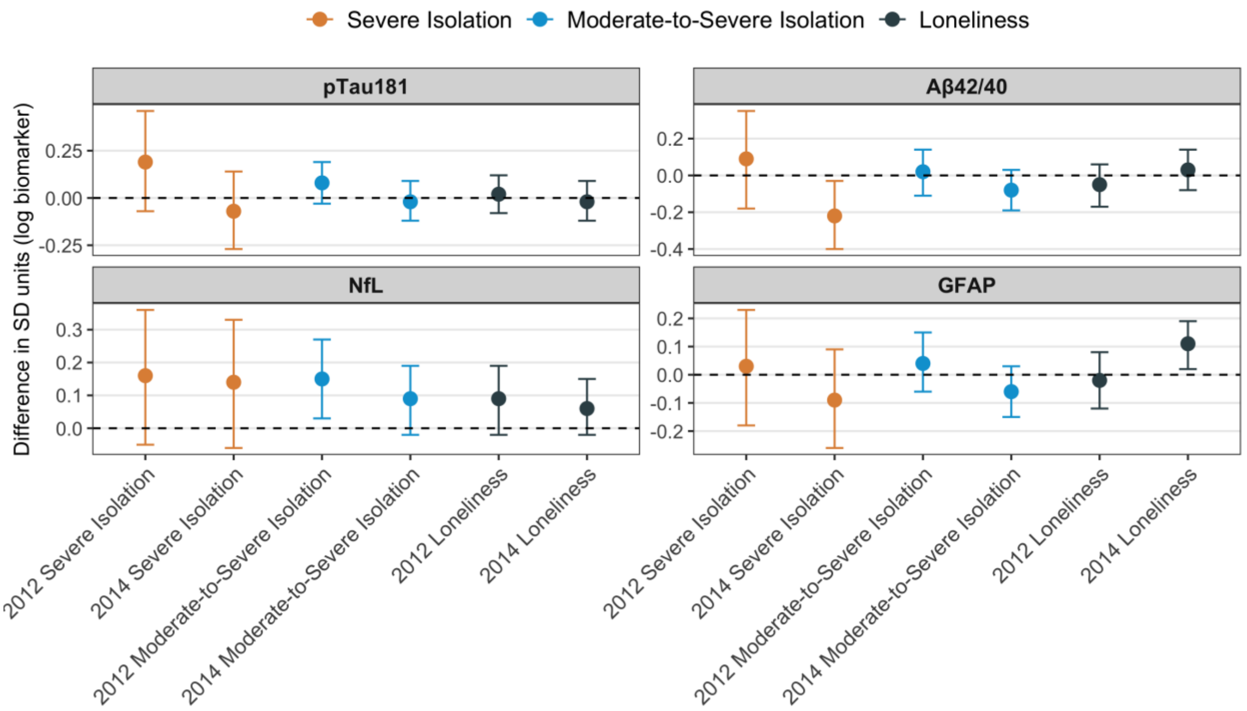

**eFigure 5. Associations between single-time-point social isolation and loneliness and blood-based AD/ADRD biomarkers in the U.S. Health and Retirement Study.**

Points represent estimated differences in standard deviation (SD) units of log-transformed biomarker levels, and error bars indicate 95% confidence intervals. Severe isolation was defined as <1 point and moderate-to-severe isolation as <2 points on an 5-point scale, with lower scores indicating more social isolation; past-week loneliness via a single-item (yes/no). Estimates are based on linear regression models with stabilized inverse probability of treatment and retention weights and robust standard errors and adjust for baseline covariates (age, sex, race/ethnicity, and educational attainment).

**eTable 17. Numerical Estimates Corresponding to eFigure 5: Associations between single-time-point social isolation and loneliness and blood-based AD/ADRD biomarkers in the U.S. Health and Retirement Study.**

| | p-tau181 | Aβ42/40 ratio | NfL | GFAP |
|---|---|---|---|---|
| **2012 Severe isolation** | 0.19 | 0.09 | 0.16 | 0.03 |
| | (-0.07 - 0.46) | (-0.18 - 0.35) | (-0.05 - 0.36) | (-0.18 - 0.23) |
| **2014 Severe isolation** | -0.07 | -0.22** | 0.14 | -0.09 |
| | (-0.27 - 0.14) | (-0.40 - -0.03) | (-0.06 - 0.33) | (-0.26 - 0.09) |
| **2012 Moderate-to-severe isolation** | 0.08 | 0.02 | 0.15** | 0.04 |
| | (-0.03 - 0.19) | (-0.11 - 0.14) | (0.03 - 0.27) | (-0.06 - 0.15) |
| **2014 Moderate-to-severe isolation** | -0.02 | -0.08 | 0.09 | -0.06 |
| | (-0.12 - 0.09) | (-0.19 - 0.03) | (-0.02 - 0.19) | (-0.15 - 0.03) |
| **2012 Loneliness** | 0.02 | -0.05 | 0.09 | -0.02 |
| | (-0.08 - 0.12) | (-0.17 - 0.06) | (-0.02 - 0.19) | (-0.12 - 0.08) |
| **2014 Loneliness** | -0.02 | 0.03 | 0.06 | 0.11** |
| | (-0.12 - 0.09) | (-0.08 - 0.14) | (-0.02 - 0.15) | (0.02 - 0.19) |

Source: Health and Retirement Study (HRS), 2010–2016. (n=3862). Severe isolation was defined as <1 point and moderate-to-severe isolation as <2 points on an 5-point scale, with lower scores indicating more social isolation; past-week loneliness via a single-item (yes/no). Estimates are based on linear regression models with stabilized inverse probability of treatment and retention weights and robust standard errors. Models adjust for baseline covariates (age, sex, race/ethnicity, and educational attainment). Biomarker outcomes are natural log-transformed. Estimates and 95% confidence intervals, shown in parentheses, represent percentage differences obtained by exponentiating model coefficients. *** p < 0.01, ** p < 0.05, * p < 0.1.
